# Integrative modelling of reported case numbers and seroprevalence reveals time-dependent test efficiency and infectious contacts

**DOI:** 10.1101/2021.10.01.21263052

**Authors:** Lorenzo Contento, Noemi Castelletti, Elba Raimúndez, Ronan Le Gleut, Yannik Schälte, Paul Stapor, Ludwig Christian Hinske, Michael Hoelscher, Andreas Wieser, Katja Radon, Christiane Fuchs, Jan Hasenauer, the KoCo19 study group

## Abstract

Mathematical models have been widely used during the ongoing SARS-CoV-2 pandemic for data interpretation, forecasting, and policy making. However, most models are based on officially reported case numbers, which depend on test availability and test strategies. The time dependence of these factors renders interpretation difficult and might even result in estimation biases.

Here, we present a computational modelling framework that allows for the integration of reported case numbers with seroprevalence estimates obtained from representative population cohorts. To account for the time dependence of infection and testing rates, we embed flexible splines in an epidemiological model. The parameters of these splines are estimated, along with the other parameters, from the available data using a Bayesian approach.

The application of this approach to the official case numbers reported for Munich (Germany) and the seroprevalence reported by the prospective COVID-19 Cohort Munich (KoCo19) provides first estimates for the time dependence of the under-reporting factor. Furthermore, we estimate how the effectiveness of non-pharmaceutical interventions and of the testing strategy evolves over time. Overall, our results show that the integration of temporally highly resolved and representative data is beneficial for accurate epidemiological analyses.

## Introduction

Social distancing, mask wearing, lockdowns and other non-pharmaceutical interventions (NPIs) are used worldwide to slow the spread of SARS-CoV-2 and to avoid overburdening health care systems. Various studies have analysed how such NPIs influence the contact rate (Latsuzbaia et al. 2020; Jarvis et al. 2020), the infection rate (Courtemanche et al. 2020; Hartl, Wäalde, and Weber 2020; Li et al. 2020; Lyu and Wehby 2020; Siedner et al. 2020), the reproduction number (Giordano et al. 2020; Zhao and Chen 2020; Liu et al. 2021; Brauner et al. 2021; Sypsa et al. 2021) and related quantities. These studies use a broad spectrum of analysis approaches, including statistical methods (e.g., generalized linear models, generalized estimating equations, machine learning) (Streeck et al. 2020; Latsuzbaia et al. 2020; Siedner et al. 2020; Courtemanche et al. 2020), compartmental models based on ordinary differential equations (ODEs) (Barbarossa et al. 2020; Jarvis et al. 2020), and agent-based models (Lorch et al. 2021), and provide important insights. However, a limitation of most studies is that they are exclusively based on official case numbers.

The officially reported case numbers provide in most countries information about the number of positive tests for viral load registered on a specific day. Such tests can either be based on the polymerase chain reaction (PCR) or on antigen detection; for brevity’s sake, in the following we will refer to them indiscriminately as diagnostic tests. However, there are several well-known issues with these numbers (Raimúndez et al. 2021). Besides reporting delays, the most important problems are that case numbers depend on the availability of tests and on the test strategy. Clearly, the number of performed tests and the selection criteria have changed over time (e.g., due to the introduction of antigen tests). As an alternative to the officially reported case numbers, the officially reported death numbers, which are generally considered as more reliable (Radon, Bakuli, et al. 2021; Pritsch et al. 2021), can be used. However, there the effects of NPIs are smoothed over time and only visible after a substantial delay. Furthermore, the observation can be confounded by the quality of medical care, in particular if the number of beds in intensive-care units becomes a limiting factor. In summary, while case and death numbers provide information on the progression of an epidemic, the interpretation is often difficult.

The ideal data-source for the analysis of NPIs as well as the efficiency of test strategies would be a thorough monitoring of a large representative population cohort. Yet this is rather time- and resource-consuming, and in most cases not realistic. Cross-sectional studies based on diagnostic tests with a high time resolution would provide a comprehensive picture of the spread within populations, but the number of required tests would be very high. For a prevalence of 100 in 100,000 individuals, on average 1000 tests have to be performed to observe a single positive individual. For tight confidence intervals, tens of thousands of diagnostic tests would be necessary per time point. To make things worse, diagnostic tests are only positive for a short period after exposure to the virus.

An alternative to diagnostic tests that allows for the monitoring of epidemics is testing for the presence of antibodies, which assesses seroprevalence. The antibody response is rather stable and can usually be detected even months after the initial infection (Radon, Bakuli, et al. 2021; Olbrich et al. 2021; Isho et al. 2020). Accordingly, antibody tests do not only provide a snapshot of the current situation as diagnostic tests do, but inform about previous exposure and hence the past history of the epidemic up to the current point. However, to provide an assessment of NPIs and test strategies with a high temporal resolution, immense resources would be required in this case too.

We believe that the most practical way to monitor at a high temporal resolution the evolution of a pandemic is to combine the frequent but biased official daily case numbers collected by the healthcare authorities with less frequent but more informative seroprevalence measurements from representative population studies. In order to prove that, in this paper we report an analysis of the first COVID-19 wave in Munich during the spring of 2020. The outline of the paper is as follows: (i) we present a compartment model for integrating officially reported case and death numbers, as well as seroprevalence data from the population-based prospective COVID-19 cohort study KoCo19 in Munich (Radon, Saathoff, et al. 2020; Pritsch et al. 2021); (ii) we fit the model with and without using seroprevalence data, showing that the additional data drastically reduces uncertainty in the hidden dynamics of the epidemic; (iii) we assess the estimates of the time-dependent effectiveness of NPIs and testing strategies, quantifying their relative contribution to the reduction in the spread of the infection.

## Results

### Compartmental model for the COVID-19 epidemic

We developed a compartmental model for the dynamics of the COVID-19 epidemic that allows for the integration of the officially reported numbers of positive diagnostic tests and COVID-19 related deaths, hospital bed usage, and seroprevalence in representative cohort studies (Figure 1). The model describes the state of individuals: susceptible, exposed, infectious, hospitalized, recovered and deceased. Several of these generic states can be further refined (Figure 1A) by distinguishing infectious individuals with and without symptoms, recovered individuals with and without detectable virus, and hospitalized individuals in ward and intensive care unit (ICU).

**Figure 1:**
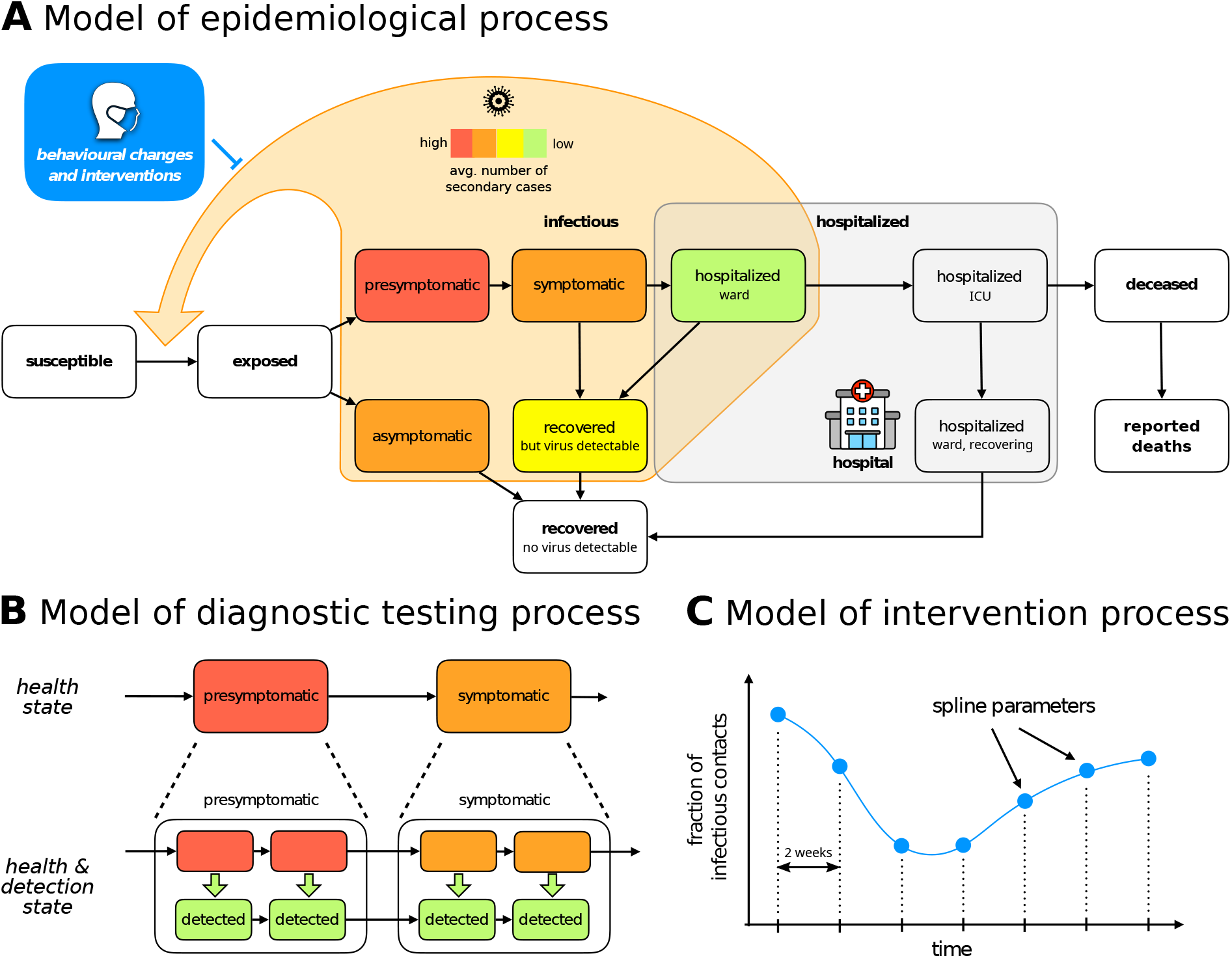
Structure of the compartment model. (A) High-level structure indicating possible transitions between various illness phases and hospitalization compartments. The delay between death and its reporting to the healthcare authorities is added in order to account for the lower number of deaths observed during weekends. Infectious phases are coloured with different colours encoding our a priori beliefs on the average number of secondary cases generated in a day by an individual in the corresponding compartment. We remark that such a number depends on the degree of infectiousness as well as the total number of inter-personal contacts. For example, on the one hand symptomatic individuals are more infectious than asymptomatic ones, but on the other they are much less likely to encounter other people due to their health condition. It is thus difficult to determine a priori which phase generates more secondary cases and as a first guess they are assigned the same colour in the figure. (B) Detailed structure of the compartment model. Each compartment is split into several sub-states in order to have Erlang-distributed transition times between compartments, and to explicitly model the testing process by tracking individuals reported to the health care authorities on a parallel but separate branch. (C) Time-dependent parameters (here the viral transmission reduction due to NPIs is used as an example) are modelled by splines which can be encoded inside the parameter vector by their values at the grid points.

Realistic distributions of transition times of individuals between states are achieved by subdividing the illness-related states into multiple sub-states (Figure 1B). This is often referred to as the Gamma Chain Trick or Linear Chain Trick (Smith 2011). Many important transition times related to the COVID-19 disease cannot be reasonably modelled by an exponential distribution (single sub-state case) — a fact that is disregarded by many studies. For example, the incubation time has been shown to be reasonably approximated by an Erlang distribution with shape parameter 6 (Lauer et al. 2020), suggesting a split into six sub-states.

The testing process is modelled by splitting illness-related states into two sub-states: infectious individuals that have not been detected, and infectious individuals that have been detected by means of a positive diagnostic test. The rates at which individuals transition from the undetected to the detected branch reflect the efficacy of the testing system set up by the healthcare authorities. As individuals with symptoms are more likely to get tested, we assume that these detection rates depend on the illness phase. In particular, the detection rate for asymptomatic and presymptomatic individuals are especially important since they can be considered a measure of contact tracing effectiveness. Hospitalized individuals are assumed to be immediately detected. The number of reported positive diagnostic tests is then equal to the sum of all fluxes from undetected to detected sub-states.

Since antibodies become detectable only about two weeks after an exposed individual has become infectious (Long et al. 2020), it is not possible to obtain the current number of seropositive individuals from the model state. Instead, we compute the number of individuals who will be seropositive in two weeks’ time, which is equal to total population size minus the number of susceptible, exposed (but not yet infectious) and deceased individuals.

The rate at which susceptible individuals are infected is proportional to the sum of the number of asymptomatic individuals, presymptomatic individuals, symptomatic individuals, recovered individuals with detectable virus levels, and hospitalized individuals in ward (ICU patients are considered to be not infectious). The elements of this sum must be weighted by the average number of secondary cases generated in a day, which is specific of each compartment. In particular, this number depends both on biological factors, such as the stage to which the illness has progressed, and behavioural factors, which in turn depend strongly on whether the infected individual has been detected or not (detected individuals are quarantined and therefore less likely to spread the disease). The qualitative ordering is indicated by colours in Figure 1A, but the quantitative contributions are considered to be unknown a priori.

### Statistical framework for inferring testing and intervention effects

The compartment model describes possible transitions between states, but the transition rates are unknown and depend on many factors. Particularly relevant is the influence on the infection and detection rates due to the policies set by the government and the healthcare authorities. We model such effects with three parameters and, since policies have evolved over time, we assume these parameters to be time-dependent, modelling them using splines (Figure 1C).

The first two are the detection rates for (i) symptomatic and (ii) asymptomatic/presymptomatic individuals respectively. These rates are influenced by testing capacity, by the effectiveness of contact tracing, and by the criteria to be met in order to be eligible for testing, all of which have undergone considerable changes in the first months of the epidemic. The third is a fractional reduction in the number of infectious contacts compared to the situation at the beginning of the epidemic. Such a reduction is strongly influenced by government restrictions such as business/school closures or lockdowns, but also depends on the compliance of the general population with social distancing and other preventive measures. In all cases time dependency is modelled by cubic splines with an inter-node distance fixed at 2 weeks. Moreover, the evolution of the detection rates has an additional week-periodic component in order to account for the lower case counts during weekends that have been observed in Germany and elsewhere in the world (see *Material and Methods* for more details). A similar week-periodic component is also present in the delay between death and its reporting to the healthcare authorities.

To determine the time-dependent infection and detection rates as well as other unknown parameters, we employ a Bayesian approach to integrate different pieces of information, including seroprevalence observed in representative cohort studies, reported case numbers, and prior knowledge on process parameters. The publicly available case counts have been modelled with negative binomial distributions, since the variance increases together with the expected value. Since hospital bed counts have a much more limited dynamic range and consequently their variance can be safely considered constant, they are assumed to be normally distributed. Finally, the number of positive antibody tests in a random sample from the population is naturally modelled as the result of a binomial random variable. The priors for the various model parameters were extracted from various published reports (see Supplementary Tables 1 and 2).

### Modelling without representative data provides uncertain estimates

As most studies are only based on officially reported case numbers, we first studied the reliability of such an approach. Therefore, we inferred the model parameters from the reported number of infected, hospitalized and deceased individuals for the city of Munich in Germany. In addition to these commonly used counts, we also employed the publicly available number of reported symptom onsets, which has been rarely used in other modelling efforts but can be easily integrated in our case thanks to our explicit description of the detection process. The number of cases (new infections, deaths and symptom onsets) was extracted from the official report by the Robert Koch Institute (2020), while the hospital usage was obtained from the web-based information system IVENA. The city of Munich was selected as detailed seroprevalence results are available from the KoCo19 study (Radon, Saathoff, et al. 2020; Pritsch et al. 2021). The time window used for this study coincides with the first COVID-19 wave in Germany, from the 1st of March to the 7th of June. The first wave is the most interesting phase for assessing the time-dependence of testing efficacy and NPIs, since several NPIs were tried in succession (up to the strictest lockdowns) and the testing capacity quickly ramped up in response to the novel virus.

Inferred parameter estimates capture correctly the case numbers used for fitting (Figure 2A). Yet, many of the estimated parameters are not well determined, as evinced by the broad credible intervals (Figure 2C, in blue). To assess the reliability of the predictions, we employed the posterior samples to predict the seroprevalence and compared it with the KoCo19 data, which was not used for fitting (Figure 2A, bottom-right panel). The prediction given by the most likely parameters encountered during sampling is compatible with the observed seroprevalances. However, as can be seen from Figure 3, the total number of cases predicted by the model has very large credible intervals. This shows that officially reported case numbers are, even in combination with prior knowledge, insufficient to predict the the actual number of COVID-19 infections during the epidemic with a satisfactory degree of confidence.

**Figure 2:**
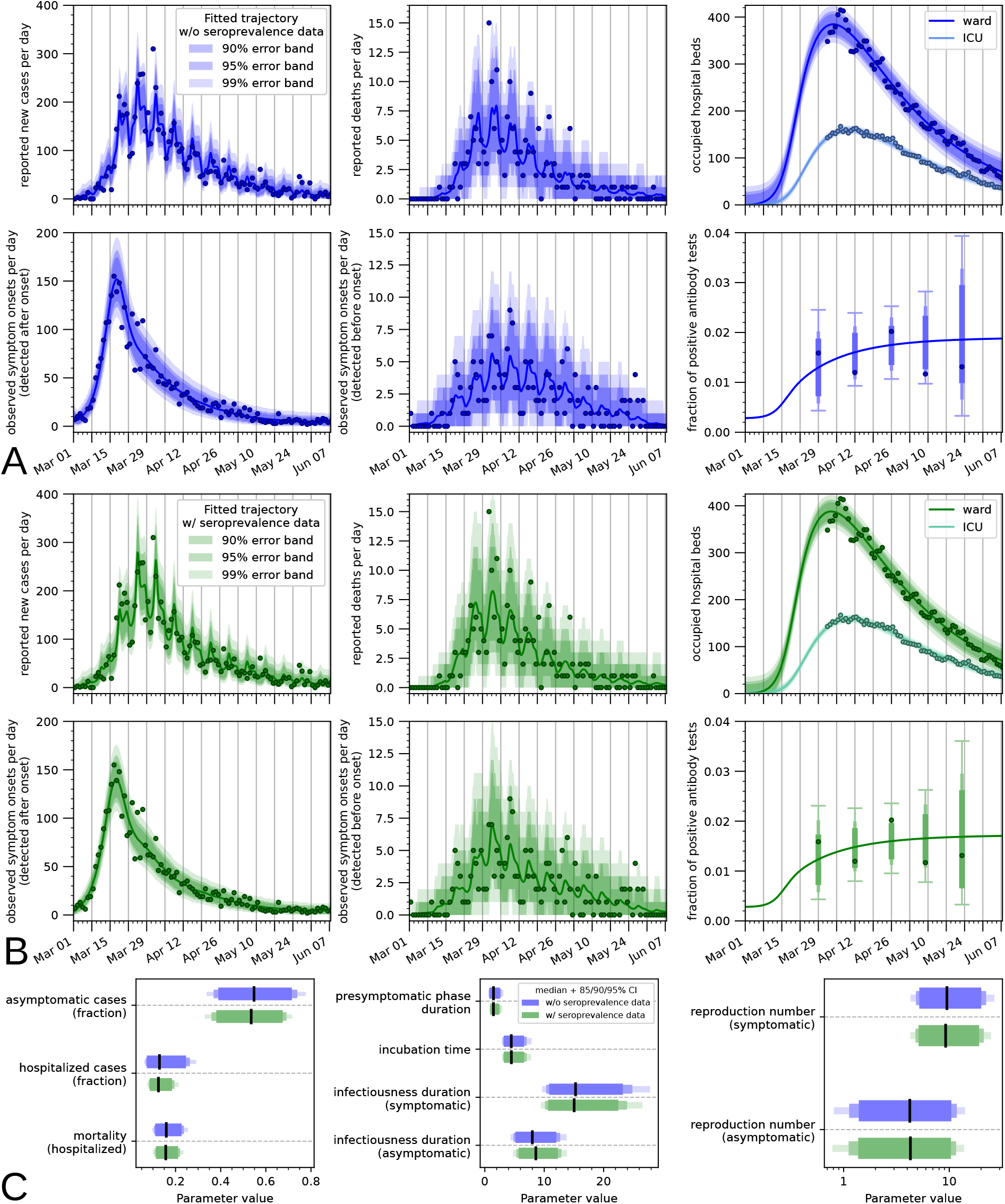
Modelling and parameter estimation for Munich, Germany. (A, B) Model simulation for the sampled parameter vector with the highest posterior probability compared with the observed data. In (A) only the case numbers reported by the Robert-Koch Institute and hospital usage for Munich are used for fitting, while in (B) seroprevalence data is also employed. The error bands show the range of plausible values for the observation, confirming that the noise models used are appropriate. In the bottom-right panels of (A, B), where the seroprevalence predicted by the model is plotted, the error bars are only shown at the observation times since the variance of each observation is linked to the number of total antibody tests carried out in each sub-batch. (C) Credible intervals (bars) and median value (line) for a subset of the model parameters. By reproduction number we mean the basic reproduction number in absence of NPIs and diagnostic testing (see *Materials & Methods* for more details on its computation).

**Figure 3:**
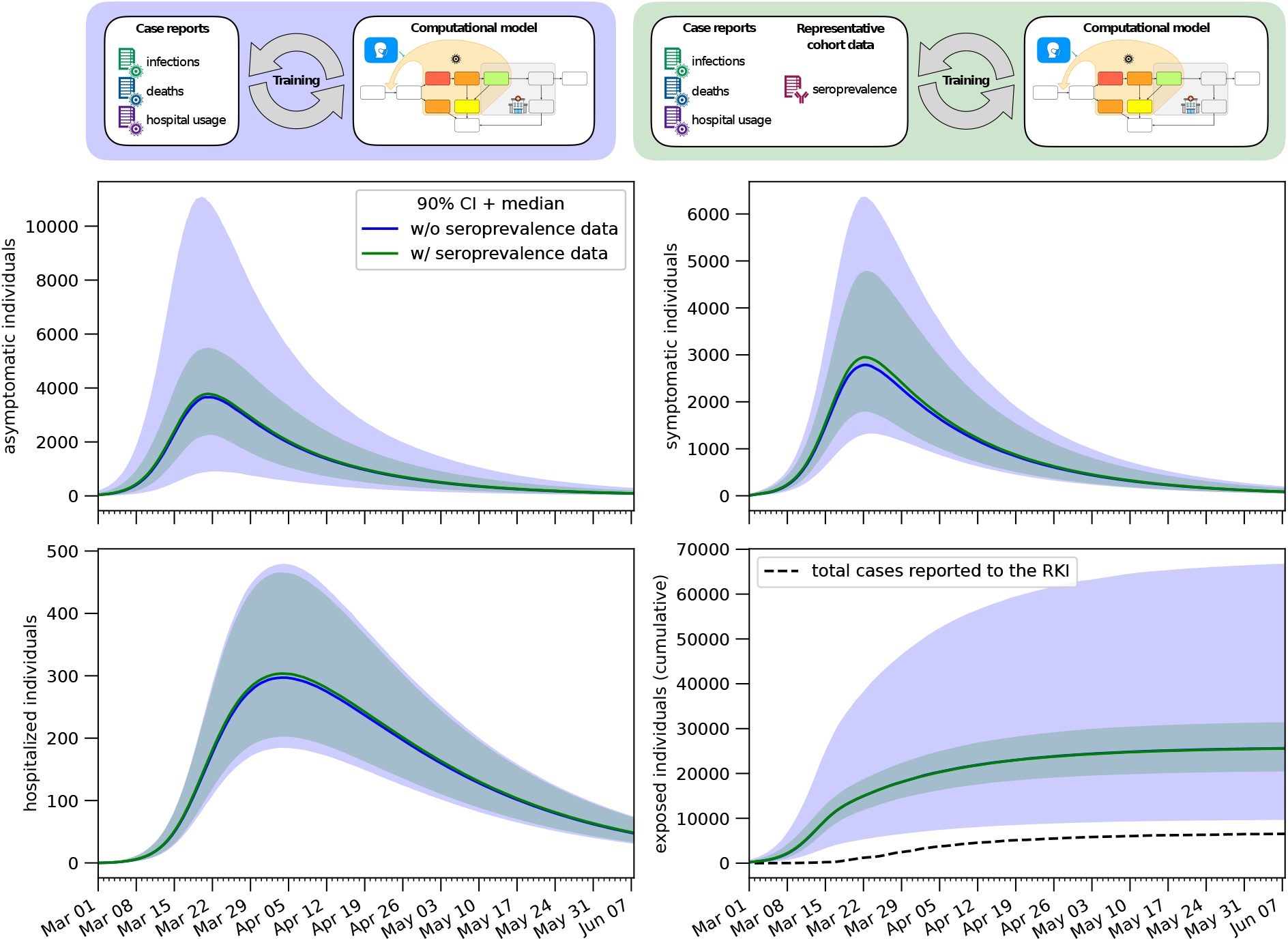
Estimates for the hidden dynamics of the epidemic. The number of individuals in different compartments is plotted as a function of time for both the model fitted with seroprevalence data and the one fitted without. In the bottom-right panel the cumulative number of cases detected by the healthcare authorities is also plotted for reference. The bands correspond to 90% posterior credible intervals, while the solid line denotes the median value.

### Modelling with representative data reduces uncertainties

In order to quantify the added value provided by prevalence data obtained by extensive serological testing, we extended the previously used dataset with the time-dependent prevalence reported by KoCo19 and performed the same Bayesian parameter estimation procedure. As in the previous analysis, the obtained parameter estimates provide an accurate description of all available data (Figure 2B) and the uncertainty on the values of the single parameters is quite large (Figure 2C, in green; for the full posterior distributions see Supplemental Figure 3). The uncertainty on the hidden states of the model (Figure 3) is instead greatly reduced, in particular for the total number of cases (not surprisingly, since this quantity is closely linked to the seroprevalence level) and for the number of asymptomatic cases, showing the effectiveness of the seroprevalence data in reducing uncertainty. Figure 3 also shows how the number of infections predicted by the model is substantially higher than the number of reported cases, highlighting the limitations of the publicly reported case counts.

### Model reveals efficiency of testing strategy

Using the compartment model integrating case reports and representative data, we studied the effectiveness of the testing and NPI strategy. To do that, we computed from the posterior samples the time-dependent detection and number of infectious contacts, along with several secondary properties.

Instead of employing directly the time-dependent detection rates, in order to evaluate the effectiveness of the testing strategy we use a more easily interpretable metric: the probability that an infected individual is reported to the healthcare authorities before the virus is cleared from their system (Figure 4B). The effectiveness of testing increased gradually as the epidemic progressed. The model estimates that at the beginning of March only 10–40% (90% CI) of infected individuals were reported, while in April and May the fraction was 25–50% (90% CI). The largest contribution is the increase in the detection probability for asymptomatic cases, which jumps from 0–10% (90% CI) to 15–40% (90% CI).

**Figure 4:**
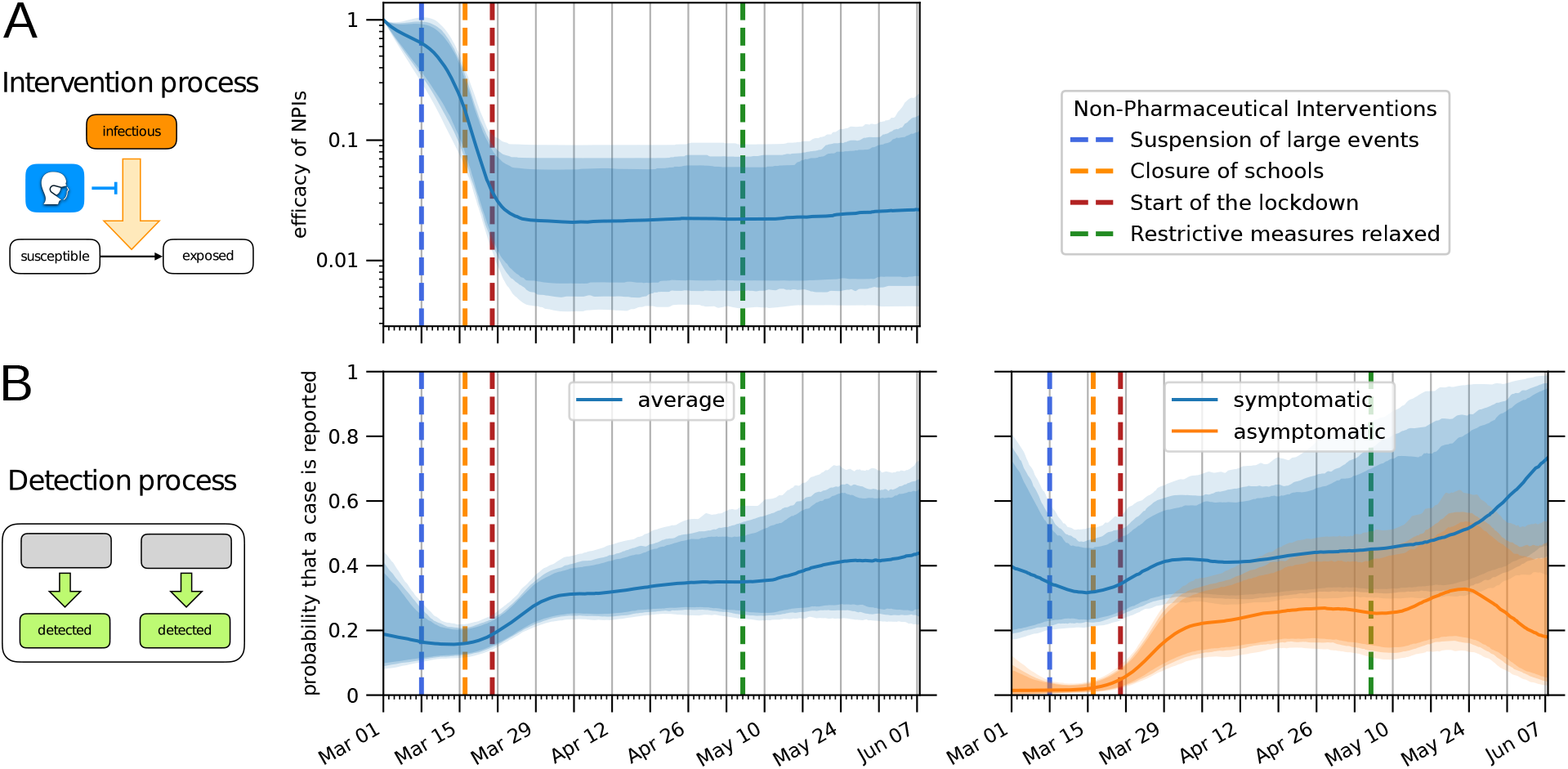
Estimation of time-dependent parameters. The bands correspond to 85%/90%/95% posterior credible intervals, while the solid line denotes the median value. Dashed lines indicate at which dates specific NPIs were enforced/lifted. (A) Reduction in the number of infectious contacts due to NPIs and behavioural changes, relative to the situation on March 1. (B) Probability that an infected individual is detected and reported to the healthcare authorities before the virus is cleared from their system. The left plot shows the average value, while the right plot shows separetely the contributions due to symptomatic and asymptomatic individuals respectively.

For the time-dependent reduction in infectious contacts, which models both the effect of government policy and behavioural changes, we observed an opposite effect compared to the detection rate (Figure 4A). Fixed to be 1 on the first day of March, this factor immediately started to drop quickly. Similarly, the effective reproduction number dropped from above three at the beginning of March to below one after the middle of March (Figure 5D). This coincides with the raising of public awareness (e.g., a speech by the German chancellor) and various interventions.

**Figure 5:**
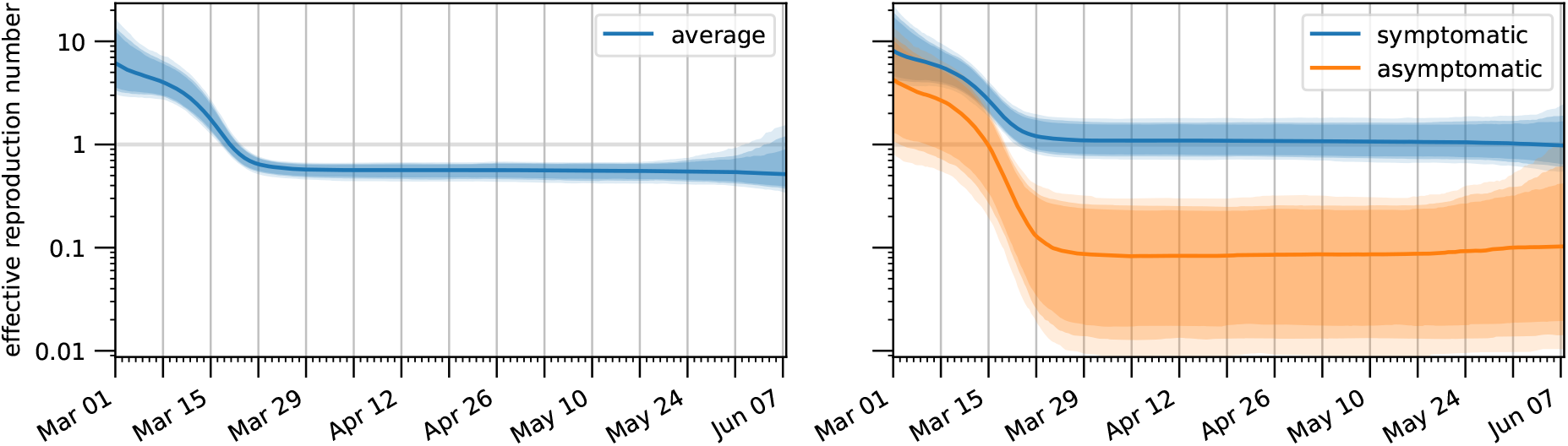
Temporal evolution of the effective reproduction number. The right plot shows the effective reproduction numbers for symptomatic and asymptomatic infected individuals, while the left plot shows their weighted average, i.e. the reproduction number for a generic infected individual. The bands correspond to 85%/90%/95% posterior credible intervals, while the solid line denotes the median value.

The effective reproduction number is influenced by both NPIs and by the testing strategy and it is therefore important to deconvolute these two effects. In order to do so, we computed the evolution of the effective reproduction number for three hypothetical scenarios (Figure 6): (i) neither NPIs are used nor diagnostic testing performed; (ii) only diagnostic testing is performed; (iii) only NPIs are employed (see *Materials & Methods* for more details on the computation). This revealed that diagnostic testing results in a small improvement over what can be achieved with NPIs alone. However, in absence of NPIs such as the lockdown, the testing strategy implemented during the first epidemic wave in Germany would not have been able to lower the reproduction number enough to stop the spreading of the disease.

**Figure 6:**
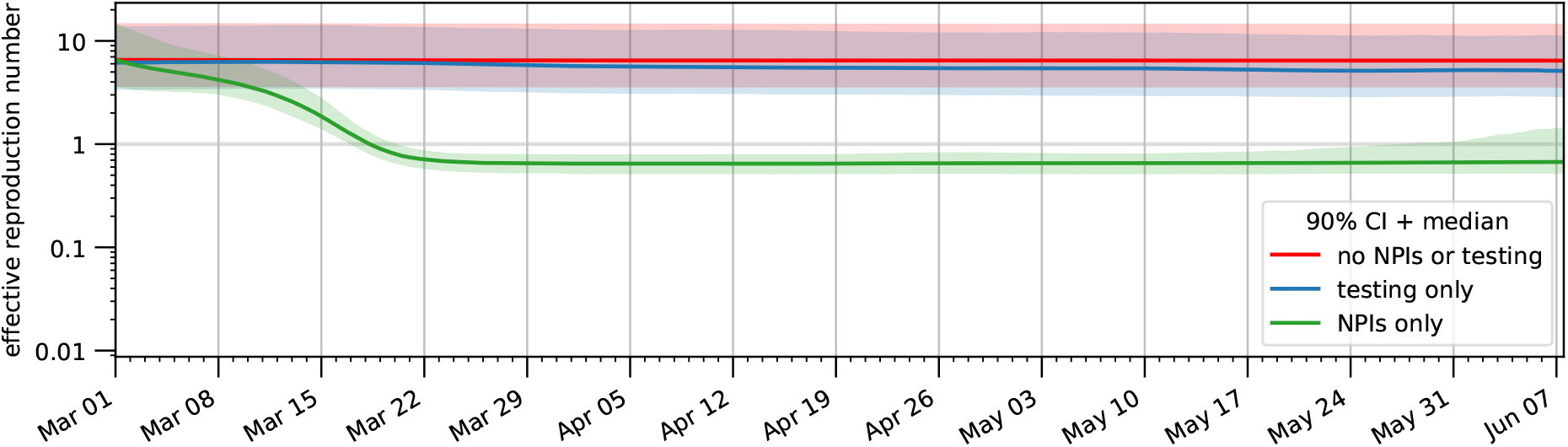
Relative importance of the detection process and the NPIs on the spread of the epidemic. Estimates for the reproduction number are plotted for three different scenarios: (red) neither diagnostic testing nor NPIs are employed; (blue) only diagnostic testing is performed; (green) only NPIs are applied. The bands correspond to the 90% posterior credible interval, while the solid line denotes the median value.

## Discussion

Test availability, testing strategy, governmental interventions and various factors have changed over the course of the COVID-19 epidemic in Germany. This renders the interpretations of the reported case numbers difficult, while creating the need to infer time-dependent characteristics (e.g., to assess the impact of strategies). Here, we approached both points by integrating officially reported case numbers with representative seroprevalence observations using integrative modelling and Bayesian parameter estimation framework. Our analysis revealed that the integration of datasets is critical: The amount of available seroprevalance data was too limited to build models just based on them, while case report numbers on their own resulted in vary large uncertainties on the hidden states of the model, especially on the number of asymptomatic cases (Figure 3). In the last year statistical works integrating case numbers and seroprevalence have been published (Quick, Dey, and X. Lin 2021), but their number is still rather limited due to the logistical difficulties of wide serological surveys.

In addition to the use of various data sources, a strength of our study is the rigorous application of Bayesian uncertainty quantification. The vast majority of models for the COVID-19 epidemic we have found in the literature were not accompanied by uncertainty quantification. Some exceptions exist, such as a study by Y. T. Lin et al. (2021), in which uncertainty estimates are given for the observed case number and the parameter values. However, they chose not to estimate most illness related parameters, but to fix them to values taken from the literature. This is problematic for two reasons: Firstly, many parameters are region- and situation-specific and can thus lead to wrong estimates of inferred parameters. Secondly, estimates typically have large uncertainty intervals, as also seen in our study and by Raimúndez et al. (2021). Fixing those to single values may lead to an underestimation of the uncertainty of inferred parameters. This was especially true during the first wave of the pandemic, when abundant observational data was still not available. For these reasons, we incorporate pre-existing knowledge from the literature only as prior information and estimate all parameters at the same time.

Our integrative modelling framework is able to estimate the effectiveness of the testing strategies employed during the first wave of the epidemic in Munich (Germany). In particular, it suggests that the fraction of detected asymptomatic SARS-CoV-2 cases quickly saturated. At the beginning of April 20% (90% CI: 10–30%) of asymptomatic individuals were detected and the numbers changed afterwards only marginally. Indeed, the model predicts even a small drop at the end of May, but the uncertainty is large (due to a low number of observations).

The proposed model relies on a detailed description of the testing process, with the inclusion of symptom onset data and time-dependent detection rates reflecting the varying test capacity of the health care system and the change of the criteria needed for obtaining a test. While still preliminary, we believe such additional modelling to be very important especially in the initial phase of an outbreak. The proposed formulation could be the basis for future studies and expanded by e.g. including age groups and interactions with neighbouring regions.

Our model is rather complex compared to what is commonly found in the literature since we have deliberately striven to model as precisely as possible the viral life cycle and the detection process. In addition to bringing us closer to the real phenomenon, a more detailed model has the advantage that model states have a more precise interpretation, allowing us to map them to a wider range of data sources, either for fitting or for obtaining prior information. This approach has however its limitations. Our complex model, coupled with our use of weak priors that reflects the lack of information at the beginning of a new pandemic, results in a very large uncertainty on most parameters, some of which are most certainly not identifiable. However, we are not mainly interested in providing tight parameter estimates but rather in assessing the added value brought by seroprevalence datasets. If one looks at the estimates of the evolution of the number of infected individuals (especially asymptomatic ones), the levels of uncertainty are much lower and the improvement due to the integration of seroprevalence data is clear. The real limitation of a complex model lies instead in the computation time it requires. A simpler model with more identifiable parameters and/or stronger priors would be more effective for time-sensitive goals, e.g. in providing online estimation and predictions for an ongoing pandemic.

Overall, the proposed work highlights the importance of seroprevalence data and thereby complements various existing efforts. We expect that it might contribute to a better understanding of the dynamics of epidemics in a dynamic environment, with changing testing capabilities and NPIs.

## Materials and Methods

### Data sources

#### Case report data

We use the official case data for Munich (Germany), which is released on a daily basis by the Robert Koch Institute (2020). We process the data to obtain the total number of new cases and deaths which were communicated to the public health officials for each day. The delay between diagnosis and recording in the RKI database can be rather long, but since we are only dealing with data from the first pandemic wave this is not a matter of concern in our study.

We can observe in the RKI dataset the so-called “weekend effect”: the number of new cases and deaths is significantly lower during the weekend. A similar effect has been observed in other countries and for other diseases as well. However, there is no consensus on the reasons behind such a periodicity. Possible explanations include: reporting delays; decreased diagnostic testing capacity; lower probability of dying during the weekend due to critical therapeutic decisions being made less often during holidays. Our model treats the weekend effect as a kind of reporting delay (see the *Detection process* paragraph for more details). Decreased testing capacity and decreased changes in therapy can also be accounted for by such an approach.

In addition to the case and death counts used in many other studies, for a subset of patients the RKI provides the date at which they first displayed symptoms. To the best of our knowledge such information has not been used in any other modelling effort. By comparing the date of symptom onset to the date of detection we can determine whether the patient was presymptomatic or symptomatic when they were tested. Since our model keeps the progression of detected and undetected individuals separate, these two counts can be fitted simultaneously.

#### Hospital bed counts

In addition to the RKI data, we employed also the number of hospital beds in Munich occupied by COVID-19 patients (IVENA). Bed counts were aggregated by hospital unit: ward, medium care unit (MCU) and intensive care unit (ICU). Since not all hospitals have a MCU, we further aggregated MCU and ICU bed counts. Due to a non-uniform reporting by the different hospitals (especially in the first days since the reporting system was set up), the data before 25 March 2020 had to be discarded. Some of the smaller hospitals/clinics did not reliably report bed counts even after this date and had to be excluded, leading to a possible, albeit slight, underestimation of the total number of occupied beds.

Another possible problem in the data is that patients may be moved from the surrounding areas to the city, where hospital concentration is higher, resulting in a possible overestimation of the number of occupied beds compared to what can be predicted by a model which does not take into account immigration effects. We thus introduce two under/over-representation factors for the bed counts, one for the occupied beds in ward and the other for the occupied ICU stations (see Supplementary Figure 7 for their posterior distributions). We have to distinguish them since the distribution of patients coming from outside the city may be skewed in favor of severe cases which cannot be treated in smaller hospitals. Evidence for this can be found in the ratio between occupied ICU and ward beds, which differs significantly when comparing the city with the whole of Bavaria (Supplementary Figure 2).

#### Seroprevalence data

We use the serological testing results reported by the “Prospective COVID-19 Cohort Munich” study (KoCo19). KoCo19 is organised by the Ludwig Maximilian University (LMU) Hospital and is currently monitoring nearly 3000 households in the Munich city area (Radon, Saathoff, et al. 2020). At regular intervals, blood samples for each household member are gathered and tested for several indicators of the presence of antibodies to SARS-CoV-2 (Olbrich et al. 2021). These tests are then used to impute the lifetime prevalence of COVID-19 in the general population (Pritsch et al. 2021), which due to the potentially large number of asymptomatic cases is impossible to recover from the case counts released by the national health authorities. In this work we employ the results from the first round of testing, spanning the period from April 6th to June 12th, 2020.

### Epidemiological model

#### Illness phases and states

At the coarsest level, individuals can be assigned to compartments corresponding to biologically different phases of the infection. From each of these phases an individual can transition to a subset of the other phases and a transition probability can be assigned to each of these possible disease progressions. Such a model can be easily visualized in graph form (see Figure 1A) and converted to a set of ODEs (mostly linear, except for the non-linear infection process). However, as mentioned in the *Results* section, transition times are usually not exponentially distributed and thus each phase is subdivided into distinct model states in order to model Erlang-distributed transition times, in what is usually referred to as the Gamma Chain Trick (Smith 2011). Additionally, the distribution of the transition times may depend not only on the current illness phase, but also on the future phase (e.g., symptomatic individuals who worsen and are hospitalized transition to the next phase faster than symptomatic individuals who are never hospitalized). When this occurs a different branch for each possible progression must be considered, each with its own transition rates and possibly with a different number of substates for the Gamma Chain Trick.

#### Detection process

As mentioned in the *Results* section, the process by which infected individuals are reported to the healthcare authorities (by means of a positive diagnostic test) is modelled explicitly with additional states that represent detected individuals. The transition rates to the model branch containing detected individuals are denoted by *k*_detect,asym_ (for asymptomatic and presymptomatic individuals) and *k*_detect,sym_ for symptomatic individuals. Since the efficacy of diagnostic testing strategies has changed during the course of the first wave of the pandemic, these two rates depend on time. We further decompose

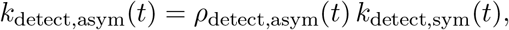

with *ρ*_detect,asym_(*t*) *∈* (0, 1), in order to encode our reasonable belief that the detection of asymptomatic individuals is more difficult than the detection of people displaying symptoms. As for the detection rate of symptomatic individuals, we write it as

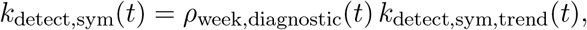

where *k*_detect,sym,trend_(*t*) *>* 0 is the long-term variation of the detection rate and *ρ*_week,diagnostic_(*t*) *>* 0 is a week-periodic term modelling the weekend effect. To reduce non-identifiability we assume that over one week *ρ*_week,diagnostic_(*t*) averages to one.

Since the weekend effect also influences death counts, a delay is also added between the time a patient dies and the time their death is reported to the healthcare authorities. This can be done by distinguishing deceased but unreported patients from reported deceased individuals, with the rate *k*_detect,death_(*t*) of the transition between these two states controlling the amount of delay present. We assume that this delay has no long-term trend and is simply given by a week-periodic function. For optimization and interpretability reasons, we decompose this value in the same way as *k*_detect,sym_, yielding

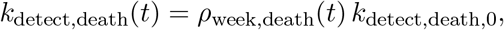

where *ρ*_week,death_(*t*) *>* 0 is a week-periodic term which averages to one over one week and *k*_detect,death,0_ is consequently the average value of the transition rate.

#### Reporting of symptom onsets

Individuals who are tested while presymptomatic and whose date of symptoms onset is reported to the healthcare authorities are those who leave the presymptomatic phase while being already detected. Individuals who are tested while symptomatic and whose date of symptoms onset is reported are instead only just a fraction of those who leave the presymptomatic phase while not being detected yet. This is due to the fact that, when the detection rate is rather low, some symptomatic individuals may not be detected before the virus is cleared from their system. The fraction of symptomatic individuals who will eventually be detected can be easily computed from the model structure (all differential equations involved are linear) and can be used to correctly scale the number of onsets. We refer to the supplemental material for the full formulas.

In order to account for the incompleteness of the symptom onset data, we introduce two scaling factors: the first ensures that already detected individuals who become symptomatic do not necessarily need to report their symptom onset date; the second allows individuals that were detected after they had developed symptoms to not report their symptom onset date. Though the uncertainty on these scaling factors is large (Supplementary Figure 7), the comparison between the posterior and prior distributions suggests that some useful information can still be extracted. We believe this also helps the model to estimate how many individuals are tested before any symptoms are observed (presumably thanks to contact tracing efforts), which in turn influences the detection rate of asymptomatic individuals.

#### Infection rate

In a simple SIR model, the equation for the change in the number of susceptible individuals *S* is given by

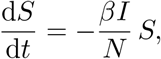

where *I* is the number of infectious individuals, *N* is the number of individuals who are alive and *β* is the (mean) number of infectious contacts an infectious individual has in a time unit, which we will assume to be equal to one day. The total number of infectious contacts occurring in a day is then *βI* and thus, assuming that such contacts are equally distributed among all alive individuals, each individual will be exposed to the virus an average of *βI/N* times per day. Since only susceptible individuals can get infected, the number of new infections per day will be equal to *βIS/N*, as in the above equation. Our model distinguishes several compartments of infectious individuals *I*_*i*_ and we denote by *β*_*i*_ the value of *β* specific to individuals in compartment *I*_*i*_. Then, the total number of infectious contacts in a day becomes 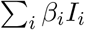 and the resulting equation is

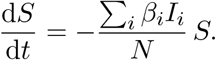

However, there are hidden dependencies between the *β*_*i*_ parameters. In order to make these relationships explicit, we have further decomposed each *β*_*i*_ as 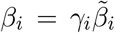, where 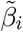 denotes the number of potentially infectious interactions that an individual from *I*_*i*_ has in a day and *γ*_*i*_ denotes the fraction of such interactions that are actually infectious. The infectiousness factor *γ*_*i*_ only depends on the biology of the disease and therefore on the illness phase. The value 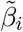 is instead a measure of the mobility/behaviour of the individuals and can depend on other factors such as the NPIs currently enforced or whether the individual has been detected by the healthcare authorities (and is thus quarantined). While different compartments usually have different values of *β*_*i*_, they may share either the value for *γ*_*i*_ (e.g., all symptomatic individuals are equally infectious, whether they have been quarantined or not) or the value for 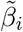 (e.g., asymptomatic individuals are thought to be less infectious than presymptomatic ones, but their social behaviour is the same since they are both unaware of their illness). This parameter sharing further constrains the model which usually has a beneficial effect on inference. Moreover, while putting priors on *β*_*i*_ would be difficult, the decomposition in biologically interpretable parameters allows us to use additional priors on *γ*_*i*_ and 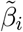 to further constrain the model to realistic parameter configurations. For example, it is expected a priori that the infectiousness *γ*_*i*_ of asymptomatic individuals should be lower than the infectiousness for symptomatic and presymptomatic individuals.

The values of 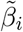 for asymptomatic and presymptomatic individuals can be considered equal since they are both unaware of their infectiousness and thus their behaviour is the same. However, since individual behaviour and mobility is affected by the NPIs currently enforced by the government, such a value must be time dependent and will be denoted by 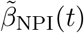. On the other hand, symptomatic individuals know they are sick (though not necessarily from COVID-19) and are limited in their mobility by their symptoms. We can safely assume that their number of potentially infectious contacts is time-independent and denote it by 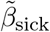. Similarly, individuals detected by diagnostic testing know they are infected and are quarantined, so that we can assign them a constant number of contacts denoted by 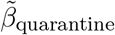 such that 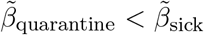. Another reasonable assumption is that 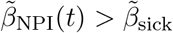. If such an assumption is not included explicitly in the model, uncertainty increases noticeably and 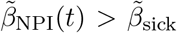 does not hold for most samples from the posterior. We thus enforce the assumption by further decomposing

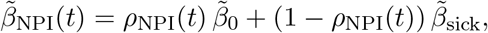

where

Where 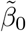 is the number of potentially infectious interactions in a pre-pandemic setting and *ρ*_NPI_(*t*) *>* 0. The choice of 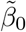 is somewhat arbitrary, since any change in it can be compensated by an opposite change in the infectiousness level *γ*_*i*_. Thus, we need to fix it (and not estimate it along with the other parameters) and as its value we choose an estimate of daily social interactions for Germany taken from the pan-European POLYMOD survey (Mossong et al. 2008). The factor *ρ*_NPI_(*t*) is instead the time-dependent reduction in potentially infectious contacts caused, e.g., by a reduction in mobility or adherence to social distancing norms, which in turn can be influenced by NPIs such as lockdowns. By definition of 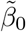, we get that *ρ*_NPI_(*t*) must be equal to 1 at the beginning of the epidemic.

The relative values for *γ*_*i*_ and *β*_*i*_ have been reported in Suppplementary Figures 4 and 5.

#### Time-dependent parameters

Time-dependent parameters are represented in our model by cubic Hermite splines defined on a grid of uniformly spaced points. The interval length has been set to a relatively large value of two weeks: this may cause some small artifacts in regions where changes occur fast, but a trade-off had to be made in order to keep computation time manageable (splines make up for the majority of model parameters). Each spline is encoded in the parameter vector by its values at grid points only and the spline derivatives are computed by finite differences, i.e., we are using a Catmull–Rom spline (Catmull and Rom 1974). A standard interpolating cubic spline would have required the same number of parameters and have been smoother (a cubic Hermite spline is only continuous up to the first derivative), but we believe the potential drawbacks were not worth it. First, a standard interpolating cubic spline requires a more difficult and computationally expensive implementation since a linear system must be solved to compute the polynomial coefficients. Second, the value of a standard spline at any given point depends on all the values at the grid points, while for a Hermite spline it depends only on the values at the two nearest grid points, preventing the unrealistic case of having parameter values at later times influence observations at earlier times (even if such influence would probably be quite small).

Our time-dependent parameters are *ρ*_NPI_, *k*_detect,sym,trend_ and *ρ*_detect,asym_ and they must be positive at all times, being transition rates or multiplicative factors. This cannot be enforced with a spline because of possible under-shooting and thus we use splines to describe the logarithm of the parameters of interest instead of the parameter values directly. As regularization, we also add zero-mean normal priors for the derivatives and the curvature of the time-dependent parameter (in its original scale, not in the logarithmic one), encoding our belief that if the data does not strongly suggest otherwise the time-dependent parameters should stay constant in value.

The two week-periodic functions *ρ*_week,diagnostic_ and *ρ*_week,death_ are (in logarithmic scale) periodic Catmull-Rom splines. In order to approximate the normalization constraint (integral over one week equal to one) imposed to ensure identifiability, by construction we force the mean of their values at the grid points (in this case, the seven days of the week) to be equal to one. As a regularization term to improve smoothness, we use the L2 norm of the derivative.

The estimate for 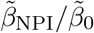 can be found in Figure 4A, the estimates for *k*_detect,sym_ and *k*_detect,asym_ are reported in Supplementary Figure 8 and the estimates for the periodic effects *ρ*_week,diagnostic_ and *ρ*_week,death_ can be found in Supplementary Figure 9.

### Observation model

Let 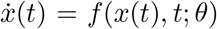be our ODE model of the epidemic, where *x* is a vector representing the model state and *θ* is the parameter vector. In most cases the model state is hidden, in the sense that it cannot be directly (or at least not exactly) measured. Let *{*(*t*_*i*_, *y*_*i*_)*}*_*i*_ be the data against which the model must be fitted. In order to link this data to the hidden state of the model, random variables *Y*_*i*_ *∼ p*_*i*_(*y*|*x*(*t*_*i*_), *θ*), known as *observable quantities*, must be introduced. Note that these observable quantities cannot be simple deterministic mappings from the state *x* to the observed values *y* since (i) measurement noise cannot be avoided; (ii) even if noise were to be eliminated, a practical model is never a perfect representation of reality. The precise distribution of the observables *Y*_*i*_ depends on the particular data source and is often referred to as the *noise model*.

#### Case counts (infections/deaths/symptom onsets reported to the RKI)

The RKI releases the total number of cases reported to the health care authorities on any given day. We compute this from the simulated model trajectory using the midpoint rule, using the instantaneous rate of detection at noon of each day. As the noise in the case counts by the RKI seems to increase as the number of cases increase (Supplementary Figure 1), a simple Gaussian noise model with constant variance is not sufficient and we use a negative binomial distribution instead. The negative binomial distribution is a generalization of the Poisson distribution which is over-dispersed, i.e., for which the *variance-to-mean ratio* (VMR, also known as *index of dispersion*) is greater than one (in the limit case in which the VMR tends to one we recover the Poisson distribution). Due to its higher flexibility the negative binomial distribution is often the preferred choice for count data (Beck and Tolnay 1995) and has been successfully employed in modelling case numbers for the COVID-19 pandemic (Y. T. Lin et al. 2021; Chan et al. 2021). We employ it too, parametrizing it by mean (the daily number of cases predicted by the model) and VMR (estimated along with the other parameters, see Supplementary Figure 6 for its posterior distribution). While our data (except for the number of new cases) is not significantly over-dispersed, we have observed the negative binomial noise model to perform much better in practice than constant-variance additive noise or log-normally-distributed multiplicative noise.

#### Hospitalization data

For the hospitalization data too it can be observed that the variance increases with the mean (Supplementary Figure 1). However, since the dynamic range of the bed counts is limited compared to the range spanned by the case counts, such an increase in variance is rather small and we can safely assume it to be approximately constant. We have found sufficient in this case to use the simpler Gaussian noise model with constant variance instead of the more complex negative binomial distribution. These variances are estimated along with the other parameters and their posterior distributions are reported in Supplementary Figure 6.

As for the time at which bed counts are observed, in this case too we assume it to be noon of each day. In this case the choice is rather arbitrary, since we have no information on when the hospitals count the number of patients and whether the methodology used is the same for all hospitals.

#### Prevalence estimates

The raw data from the serological testing study consists in a test date and test result for each participant. The period corresponding to the first round of testing is split into equal-duration intervals and the antibody test results are aggregated accordingly. Assuming that each of the resulting population subsets is representative of the whole, then the number of positive tests in each interval is a realization of a binomial random variable, with success probability equal to the prevalence (after correcting for the sensitivity and specificity of the test used) and number of trials equal to the total number of tests in that batch.

### Bayesian parameter estimation and priors

Our approach to parameter inference is fully Bayesian: we are interested in sampling probable parameter values according to the posterior distribution

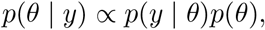

in which *p*(*y* | *θ*) is the likelihood of the observed data and *p*(*θ*) is a prior distribution on the parameter values. The likelihood can be computed from the probability distributions of the observable quantities as

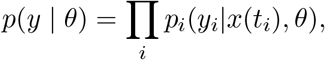

so that only the prior distribution is left to be defined.

The prior distribution should encode all available information about the parameters not coming directly from the data against which the model is fitted. In our case such information is mainly derived from clinical cases reported in the pre-existing literature. For example, the distribution of the transition times between different illness phases (e.g., incubation time) has been the object of many publications and can be used to obtain a reasonable value for the number of stages in the Gamma Chain Trick expansion. Once the number of stages is set, the same transition time distribution leads to a prior on the transition rates. Often priors can be obtained both for single parameters and for more complex expressions containing several of them. For example, in the case of transition times some overlap may be present: in one source information about the total hospitalization time may be given, in another the distribution of the time spent in the ICU may be presented and in a third the time from symptom onset to death (which in our model must necessarily transit through the ICU compartment) may be described. We have found that adding priors to derived expressions helps to exclude unrealistic values that would not have been excluded by simply putting priors on the basic parameters.

We will now show how we deal with priors on multiple expressions which are not independent. We start from the simple case where we have prior information on two quantities that can be expressed as *g*_1_(*θ*) and *g*_2_(*θ*). More than one parameter may contribute to both expressions, meaning that, even if the components of *θ* are independent, *g*_1_ and *g*_2_ may not be independent. The prior information for these two quantities is encoded in two probability distributions *p*_1_(*g*_1_) and *p*_2_(*g*_2_). One could try to construct a prior *p*(*θ*) for the full parameter vector such that the marginal distributions for the expressions *g*_1_(*θ*) and *g*_2_(*θ*) are exactly *p*_1_(*g*_1_) and *p*_2_(*g*_2_). However, such distribution may not exist (since *p*_1_ and *p*_2_ may be based on different studies and be even slightly incompatible), if it exists it may be not unique and possible solutions can only be obtained by numerical approximation. Instead we define the prior as proportional to the product *p*_1_(*g*_1_(*θ*)) *p*_2_(*g*_2_(*θ*)). The advantage of this choice for the prior is that it has an analytical formula. However, the marginal distributions for the two expressions are not *p*_1_ and *p*_2_. The posterior distribution for *g*_1_(*θ*) is not only influenced by the explicit contribution of the term *p*_1_(*g*_1_(*θ*)) but also by the implicit contribution of *p*_2_(*g*_2_(*θ*)), since some parameters appear in both expressions. In general, given an arbitrary number of expressions *g*_*k*_ for which we have prior information *p*_*k*_ as a probability distribution, we will use the non-normalized prior distribution

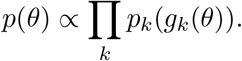

Computation of the normalization coefficient is not necessary in order to draw from the posterior distribution using MCMC. The shifting of the marginal for *g*_*k*_ from the explicit contribution *p*_*k*_ may not be a big problem since they are due to the prior taking into account all available evidence.

A summary of literature-derived priors used in our model is reported in Supplementary Tables 1 and 2. We have seen that choosing the appropriate source for prior information is difficult. For example, while the biology of the virus is constant across all countries (at least during the first wave of the epidemic, before the rise of any variant) other processes such as infection and hospitalization are extremely country specific. For this reason, we have given more weight to reports on German cases over international studies or meta-analyses. A second problem is that there is no standardization in the medical literature regarding what are the quantities of interest that should be described. For example, mean hospitalization times are often reported separately for patients that need ICU care, patients that are ventilated, patients with severe or mild symptoms, or other subsets of patients. In the worst cases the explanations provided are insufficient to exactly determine what was actually measured, making it difficult if not impossible to map the information to the model. For example, hospitalization can be considered to end with death or discharge or both, but not all sources are clear on which definition they use. Additionally, incomplete information is often reported, such as only giving median transition times instead of more accurate statistics, such as interquartile ranges. A third problem is that often prior information obtained from different sources may appear to be somehow contradictory, a problem which is obviously exacerbated by the previous point. We have tried to only include prior information of whose meaning we were quite confident but still some slight incongruities could not be removed. We opted to employ particularly weakened priors in such cases rather than completely throw the information away. A final problem is that unless the reported values are stratified by age group they may not be applicable to different waves of the epidemic in which the affected demographics differ (e.g., fraction of hospitalized individuals). In our case this is not problematic since we have only modelled the first wave of the epidemic using prior information from studies conducted on patients from the same first wave.

### Computational modelling pipeline

We established a reusable computation pipeline to automate the fitting process. The compartment model is encoded in the Systems Biology Markup Language (SBML, Hucka et al. (2003)), a widely used standard in the systems biology community. The datasets used for fitting, together with parameter priors and bounds, are stored in the PEtab format (Schmiester et al. 2021), a standard format for the formulation of estimation problems. Both formats are supported by various simulation and analysis tools, which aids accessibility and reusability of model and results.

Using the SBML models and the data in PEtab format, we perform numerical simulation and sensitivity calculation with the C++/Python library AMICI (Fröhlich et al. 2021), and parameter estimation using the Python-based tool pyPESTO (Schälte et al. 2021). This combination of tools offers a broad spectrum of functionalities, including advanced gradient-based nonlinear optimization, profile calculation, sampling and ensemble uncertainty analysis. In the current phase, we are using the optimization and the sampling capabilities to infer the unknown process parameters, including the effects of different interventions.

For sampling from the posterior distribution we use the pyPESTO interface to the state-of-theart gradient-based No-U-Turn sampler (NUTS, Hoffman and Gelman (2014)) implemented in the library PyMC3 (Salvatier, Wiecki, and Fonnesbeck 2016). As parameter estimation with a high number of parameters (75 in our case) is computationally extremely demanding, we employed parallelization on a local high-performance cluster. Yet, as sampling by the NUTS sampler is intrinsically a sequential process, each individual chain can still take weeks before producing a sufficiently high number of samples. In particular, for the results used in the paper we drew 7200 samples (excluding 2100 tuning samples) which took 2 months of wall time with a Intel^®^ Xeon^®^ Gold 6130 CPU @ 2.10GHz. Even with half the amount of samples, convergence metrics (R-hat and Geweke tests) and visual inspection of the chains were already acceptable. Effective sample size (ESS) for some parameters remains comparatively low, but this is not unexpected for a model with such a large number of parameters.

To facilitate reuse and extension, we openly share the analysis tools with the community^1^.

### Computing the reproduction number

The basic reproduction number *R*_0_ is the average number of secondary cases (new infections) generated in a completely susceptible population by a single primary case. *R*_0_ can be computed as

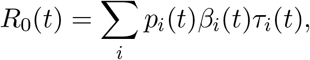

where *p*_*i*_ is the probability of an exposed individual of transiting through state *I*_*i*_, *β*_*i*_ is the average number of new cases generated by an infected individual from state *I*_*i*_ in a day and *τ*_*i*_ is the average time an individual in state *I*_*i*_ takes to transition to the next state. Since all equations are linear (except the terms dealing with infection which are not relevant to the calculation of *R*_0_), all of the three quantities can be computed analytically from the reaction rates. Moreover, since all three of them depend on time, *R*_0_ depends on time too. Finally, we want to point out that the above formula is not strictly correct, since as an individual progress through the illness states also time progresses, which would require using different values of *t* in the different states *I*_*i*_. However, since the temporal scale at which *p*_*i*_, *β*_*i*_ and *τ*_*i*_ change (which is dictated by government policies, behavioural shifts and biological evolution of the virus) is rather longer than the time required for the illness to run its course, the proposed formula is a very good approximation of the true reproduction number.

The effective reproduction number *R*_*e*_ is the average number of secondary cases generated by a primary case in a partially susceptible population. It can be easily computed from *R*_0_ as *R*_*e*_(*t*) = *R*_0_(*t*)*S*(*t*)*/N*. We can also compute a separate reproduction number *R*_0,sym_ (respectively, *R*_0,asym_) for symptomatic (respectively, asymptomatic) individuals, defined as the average number of secondary cases generated in a completely susceptible population by a primary case assuming they will (respectively, will not) develop symptoms. The formula to be used is essentially the same, with the only difference being the probabilities *p*_*i*_ have to be conditioned on the individual eventually developing (respectively, never developing) symptoms. It also holds that *R*_0_(*t*) = *f*_asym_ *R*_0,asym_ +(1*−f*_asym_) *R*_0,asym_, where *f*_asym_ is the fraction of individuals who will never develop symptoms.

We also compute variants of the reproduction number in the case where NPIs and/or diagnostic testing are removed (see Figure 6). More precisely, NPIs can be “switched off” by fixing *ρ*_NPI_(*t*) to be constantly equal to one, while diagnostic testing can be eliminated by setting all detection rates to zero. In order to estimate the base infectiousness of the virus (as in the credible intervals of Figure 2C and in the posterior distributions of Supplemental Figure 3) we use the basic reproduction number in the case where both NPIs and diagnostic testing are absent: having no NPIs in place puts us in a pre-pandemic setting, while removing diagnostic testing removes the additional uncertainty coming from the estimates of the detection rates.

## Supporting information

Model description and equations

## Data Availability

The data used in this manuscript (except data from public sources such as the Robert Koch Institute) cannot be made public due to patient consent.

## Funding

This study was funded by the Bavarian State Ministry of Science and the Arts, the University Hospital of Ludwig-Maximilians-University Munich, the Helmholtz Centre Munich, the University of Bonn (via the Transdiciplinary Research Areas), the University of Bielefeld, Munich Center of Health (McHealth) and the German Ministry for Education and Research (MoKoCo19, reference number 01KI20271), German Research Foundation (SEPAN, reference number HA 7376/3-1), Volkswagen Stiftung (reference number: 99 450). This work was supported by the Deutsche Forschungsgemeinschaft (DFG, German Research Foundation) under Germany’s Excellence Strategy EXC 2047/1 - 390685813 and EXC 2151 - 390873048. The ORCHESTRA project has received funding from the European Union’s Horizon 2020 research and innovation programme under grant agreement No 101016167. The views expressed in this paper are the sole responsibility of the authors and the Commission is not responsible for any use that may be made of the information it contains. The funders had no role in study design, data collection, data analyses, data interpretation, writing, or submission of this manuscript.

## Acknowledgments

We gratefully thank all participants of the KoCo19 study for their trust, time, data, and specimens. This study would also not have been possible without the staff of the Division of Infectious Diseases and Tropical Medicine at the University Hospital of LMU Munich, Helmholtz Centre Munich, and Bundeswehr Institute of Microbiology, as well as all medical students involved. We thank the KoCo19 advisory board members Stefan Endres, Stephanie Jacobs, Bernhard Liebl, Michael Mihatsch, Matthias Tschöp, Manfred Wildner, and Andreas Zapf.

## Declaration of Competing Interest

The authors declare that they have no known competing financial interests or personal relationships that could have appeared to influence the work reported in this paper.

## Institutional Review Board Statement

The study protocol was approved by the Institutional Review Board at the Ludwig-MaximiliansUniversity in Munich, Germany (opinion date 31 March 2020, number 20-275, opinion date amendment: 10 October 2020), prior to study initiation.

## Notes

### Competing Interest Statement

The authors have declared no competing interest.

### Author Declarations

The study protocol was approved by the Institutional Review Board at the Ludwig-Maximilians-University in Munich, Germany (opinion date 31 March 2020, number 20-275, opinion date amendment: 10 October 2020), prior to study initiation.

### Summary of Updates

New simulations data with improved prior distribution (qualitative results are unchanged). Materials and Methods section has been enlarged to include more details about the modelling. Additional figures both in text and supplement have been added. Additional supplement with full model equations has been added.

## References

Barbarossa, Maria Vittoria et al. (2020). “Modeling the spread of COVID-19 in Germany: Early assessment and possible scenarios”. In: PLOS ONE 15.9, e0238559. DOI: 10.1371/journal.pone.0238559.

Beck, E. M. and Stewart E. Tolnay (1995). “Analyzing Historical Count Data”. In: Historical Methods 28.3, pp. 125–131. DOI: 10.1080/01615440.1995.9956360.

Brauner, Jan M. et al. (2021). “Inferring the effectiveness of government interventions against COVID-19”. In: Science 371.6531. DOI: 10.1126/science.abd9338.

Catmull, Edwin and Raphael Rom (1974). “A class of local interpolating splines”. In: Computer Aided Geometric Design. Ed. by Robert E. Barnhill and Richard F. Riesenfeld. Academic Press, pp. 317–326. ISBN: 978-0-12-079050-0. DOI: 10.1016/B978-0-12-079050-0.50020-5.

Chan, Stephen et al. (2021). “Count regression models for COVID-19”. In: Physica A 563, p. 125460. DOI: 10.1016/j.physa.2020.125460.

Courtemanche, Charles et al. (2020). “Strong Social Distancing Measures In The United States Reduced The COVID-19 Growth Rate”. In: Health Affairs 39.7, pp. 1237–1246. DOI: 10.1377/hlthaff.2020.00608.

Fröhlich, Fabian et al. (Apr. 2021). “AMICI: high-performance sensitivity analysis for large ordinary differential equation models”. In: Bioinformatics. btab227. DOI: 10.1093/bioinformatics/btab227.

Giordano, Giulia et al. (June 2020). “Modelling the COVID-19 epidemic and implementation of population-wide interventions in Italy”. en. In: Nature Medicine 26.6, pp. 855–860. DOI: 10.1038/s41591-020-0883-7.

Hartl, Tobias, Klaus Wälde, and Enzo Weber (2020). “Measuring the impact of the German public shutdown on the spread of Covid-19”. In: Covid Economics 1, pp. 25–32.

Hoffman, Matthew D and Andrew Gelman (2014). “The No-U-turn sampler: Adaptively setting path lengths in Hamiltonian Monte Carlo.” In: Journal of Machine Learning Research 15.1, pp. 1593–1623.

Hucka, M. et al. (2003). “The systems biology markup language (SBML): A medium for representation and exchange of biochemical network models”. In: Bioinformatics 19.4, pp. 524–531. DOI: 10.1093/bioinformatics/btg015.

Isho, Baweleta et al. (2020). “Persistence of serum and saliva antibody responses to SARS-CoV-2 spike antigens in COVID-19 patients”. In: Science Immunology 5.52, eabe5511. DOI: 10.1126/sciimmunol.abe5511.

IVENA eHealth — interdisziplinärer Versorgungsnachweis (n.d.). Accessed: 2021-09-02. URL: https://www.ivena.de.

Jarvis, Christopher I et al. (2020). “Quantifying the impact of physical distance measures on the transmission of COVID-19 in the UK”. In: BMC Medicine 18.124. DOI: 10.1186/s12916-020-01597-8.

Latsuzbaia, Ardashel et al. (Aug. 2020). “Evolving social contact patterns during the COVID-19 crisis in Luxembourg”. In: PLOS ONE 15.8, pp. 1–13. DOI: 10.1371/journal.pone.0237128.

Lauer, Stephen A. et al. (May 5, 2020). “The Incubation Period of Coronavirus Disease 2019 (COVID-19) From Publicly Reported Confirmed Cases: Estimation and Application”. In: Annals of Internal Medicine 172.9, pp. 577–582. DOI: 10.7326/M20-0504.

Li, Michael Lingzhi et al. (2020). “Forecasting COVID-19 and Analyzing the Effect of Government Interventions”. In: medRxiv. DOI: 10.1101/2020.06.23.20138693.

Lin, Yen Ting et al. (Jan. 2021). “Daily Forecasting of New Cases for Regional Epidemics of Coronavirus Disease 2019 with Bayesian Uncertainty Quantification”. In: medRxiv. DOI: 10.1101/2020.07.20.20151506.

Liu, Y et al. (2021). “The impact of non-pharmaceutical interventions on SARS-CoV-2 transmission across 130 countries and territories”. In: BMC Med 19, p. 40. DOI: 10.1186/s12916-020-01872-8.

Long, Quan-Xin et al. (2020). “Antibody responses to SARS-CoV-2 in patients with COVID-19”. In: Nature Medicine 26, pp. 845–848. DOI: 10.1038/s41591-020-0897-1.

Lorch, Lars et al. (2021). Quantifying the Effects of Contact Tracing, Testing, and Containment Measures in the Presence of Infection Hotspots. arXiv: 2004.07641 [cs.LG].

Lyu, Wei and George L Wehby (2020). “Community Use Of Face Masks And COVID-19: Evidence From A Natural Experiment Of State Mandates In The US”. In: Health Affairs 39.8, pp. 1419– 1425. DOI: 10.1377/hlthaff.2020.00818.

Mossong, Jöel et al. (2008). “Social Contacts and Mixing Patterns Relevant to the Spread of Infectious Diseases”. In: PLOS Medicine 5.3, e74. DOI: 10.1371/journal.pmed.0050074.

Olbrich, Laura et al. (2021). “A Serology Strategy for Epidemiological Studies Based on the Comparison of the Performance of Seven Different Test Systems - The Representative COVID-19 Cohort Munich”. In: medRxiv. DOI: 10.1101/2021.01.13.21249735.

Pritsch, Michael et al. (2021). “Prevalence and Risk Factors of Infection in the Representative COVID-19 Cohort Munich”. In: International Journal of Environmental Research and Public Health 18.7, p. 3572. DOI: 10.3390/ijerph18073572.

Quick, Corbin, Rounak Dey, and Xihong Lin (2021). “Regression Models for Understanding COVID-19 Epidemic Dynamics With Incomplete Data”. In: Journal of the American Statistical Association 116.536, pp. 1561–1577. DOI: 10.1080/01621459.2021.2001339.

Radon, Katja, Abhishek Bakuli, et al. (2021). “From first to second wave: follow-up of the prospective Covid-19 cohort (KoCo19) in Munich (Germany)”. In: medRxiv. DOI: 10.1101/2021.04.27.21256133.

Radon, Katja, Elmar Saathoff, et al. (June 2020). “Protocol of a population-based prospective COVID-19 cohort study Munich, Germany (KoCo19)”. In: BMC Public Health 20 (1), p. 1036. DOI: 10.1186/s12889-020-09164-9.

Raimúndez, Elba et al. (2021). “COVID-19 outbreak in Wuhan demonstrates the limitations of publicly available case numbers for epidemiological modeling”. In: Epidemics 34, p. 100439. DOI: https://doi.org/10.1016/j.epidem.2021.100439.

Robert Koch Institute (2020). COVID-19: Fallzahlen in Deutschland. URL: https://npgeo-corona-npgeo-de.hub.arcgis.com/datasets/dd4580c810204019a7b8eb3e0b329dd6_0.

Salvatier, John, Thomas V. Wiecki, and Christopher Fonnesbeck (Apr. 2016). “Probabilistic programming in Python using PyMC3”. In: PeerJ Computer Science 2016 (4), e55. DOI: 10.7717/peerj-cs.55. URL: https://github.com/pymc-devs/pymc3.

Schälte, Yannik et al. (2021). ICB-DCM/pyPESTO: pyPESTO 0.2.5. DOI: 10.5281/zenodo.2553546.

Schmiester, Leonard et al. (Jan. 2021). “PEtab—Interoperable specification of parameter estimation problems in systems biology”. In: PLOS Computational Biology 17.1, pp. 1–10. DOI: 10.1371/journal.pcbi.1008646.

Siedner, Mark J. et al. (Aug. 2020). “Social distancing to slow the US COVID-19 epidemic: Longitudinal pretest–posttest comparison group study”. In: PLOS Medicine 17.8, pp. 1–12. DOI: 10.1371/journal.pmed.1003244.

Smith, Hal (2011). “Distributed Delay Equations and the Linear Chain Trick”. In: Springer, New York, NY, pp. 119–130. DOI: 10.1007/978-1-4419-7646-8_7.

Streeck, Hendrik et al. (2020). “Infection fatality rate of SARS-CoV2 in a super-spreading event in Germany”. In: Nature Communications 11.1, pp. 1–12.

Sypsa, V et al. (2021). “Effects of Social Distancing Measures during the First Epidemic Wave of Severe Acute Respiratory Syndrome Infection, Greece”. In: Emerging Infectious Diseases 27.2, pp. 452–462. DOI: 10.3201/eid2702.203412.

Zhao, S and H Chen (2020). “Modeling the epidemic dynamics and control of COVID-19 outbreak in China”. In: Quant Biol 8, pp. 11–19. DOI: 10.1007/s40484-020-0199-0.

