## Supplementary figures and tables for "Integrative modelling of reported case numbers and seroprevalence reveals time-dependent test efficiency and infectious contacts"

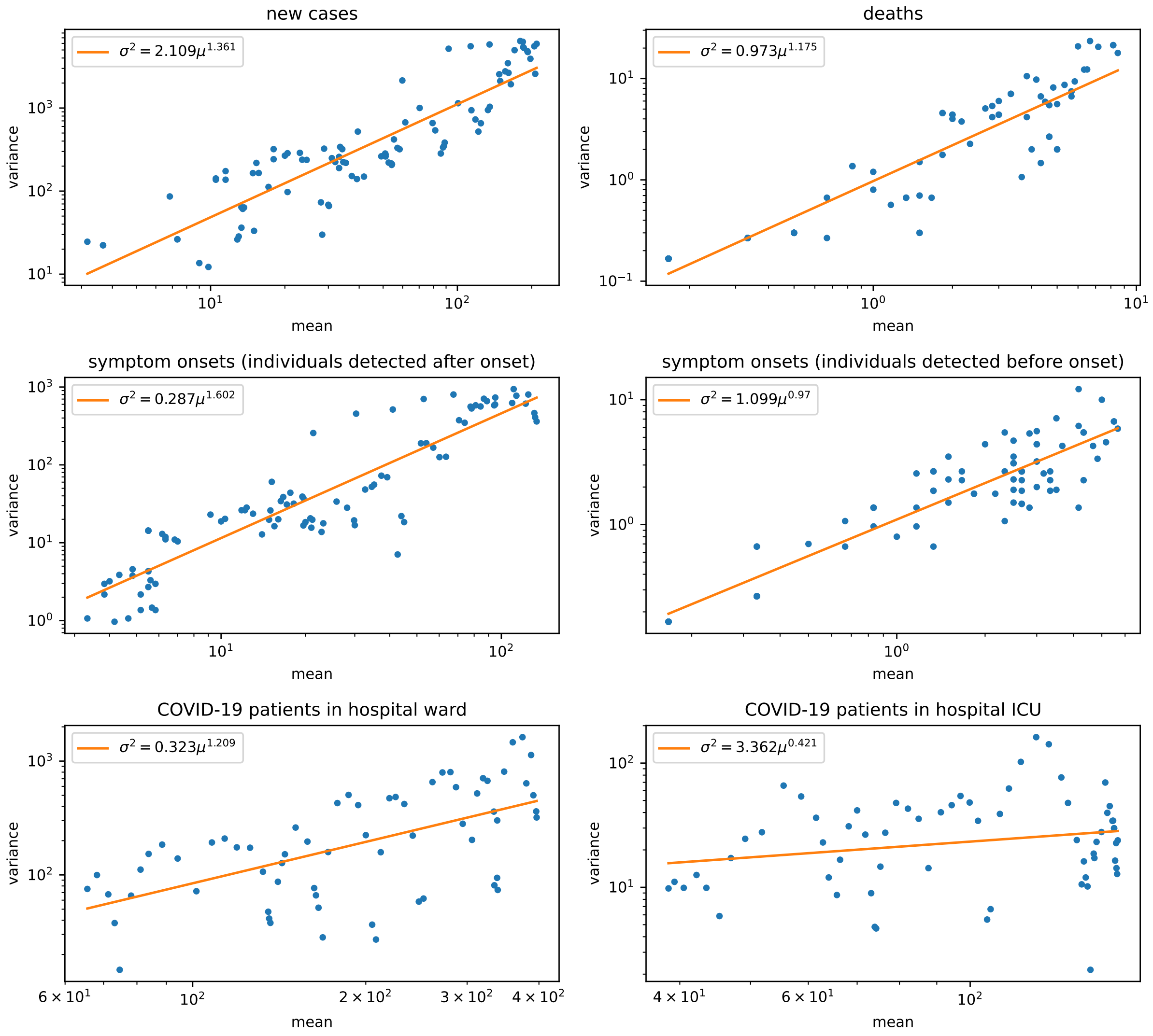


**Supplementary Figure 1: Relation between mean and variance in case count data**. Means and variances are computed from the timeseries over windows 5 days long.

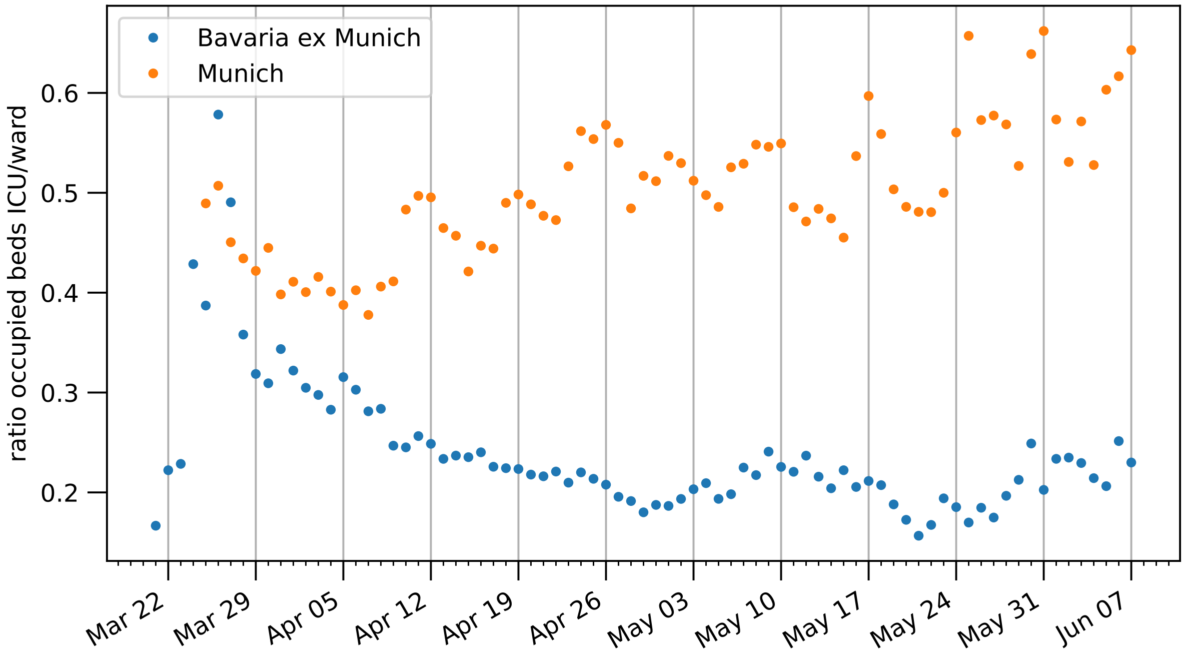


**Supplementary Figure 2:** Ratio between beds occupied by COVID-19 patients in ICU and ward, respectively for the city of Munich and the rest of Bavaria.

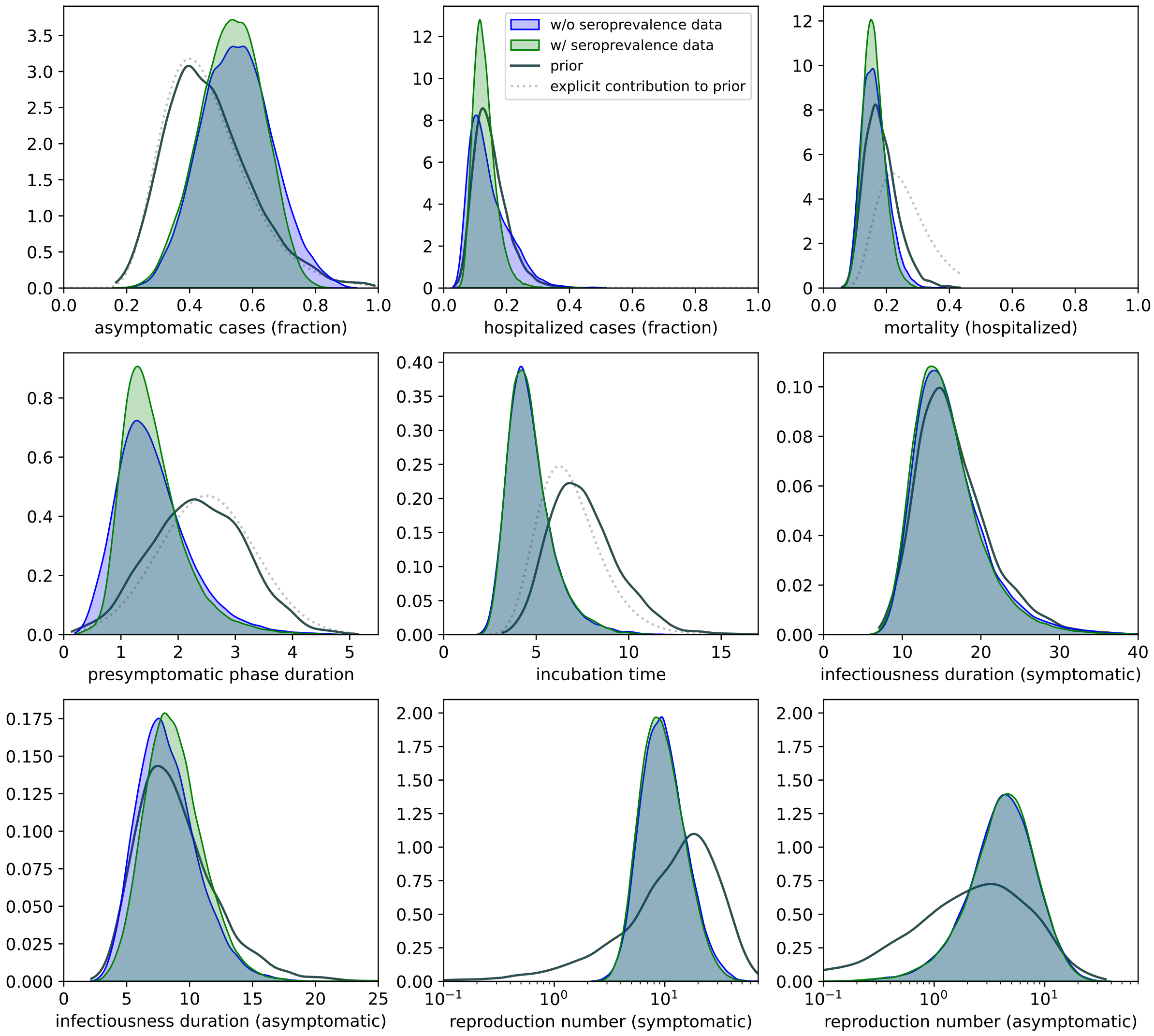


**Supplementary Figure 3: Posterior distributions for a small subset of significant model parameters.** We have plotted the KDEs obtained for both the case in which seroprevalance data is used for fitting and the one in which it is not. Prior distributions are also plotted: the solid lines are the KDEs obtained from MCMC sampling of the prior, while the dotted lines (where present) are the contributions to the prior density due to the quantity under consideration.

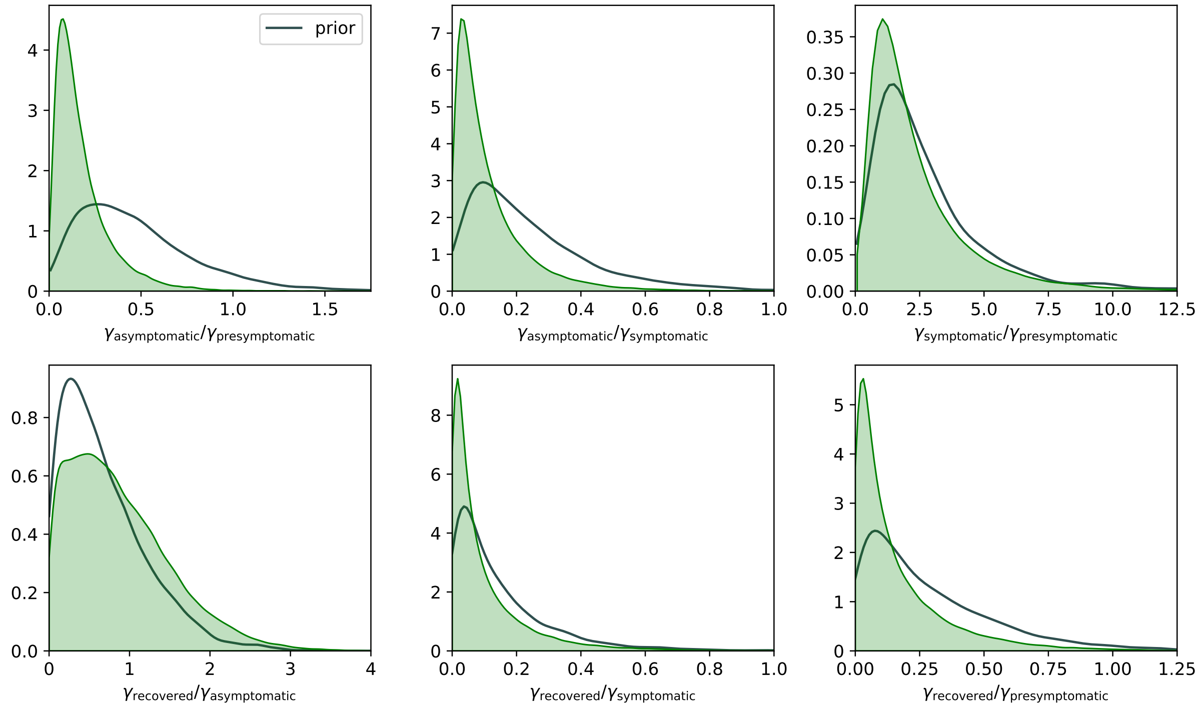


**Supplementary Figure 4: Posterior distributions for the infectiousness level $\gamma$.** Only the prior and the results obtained using seroprevalence data have been plotted.

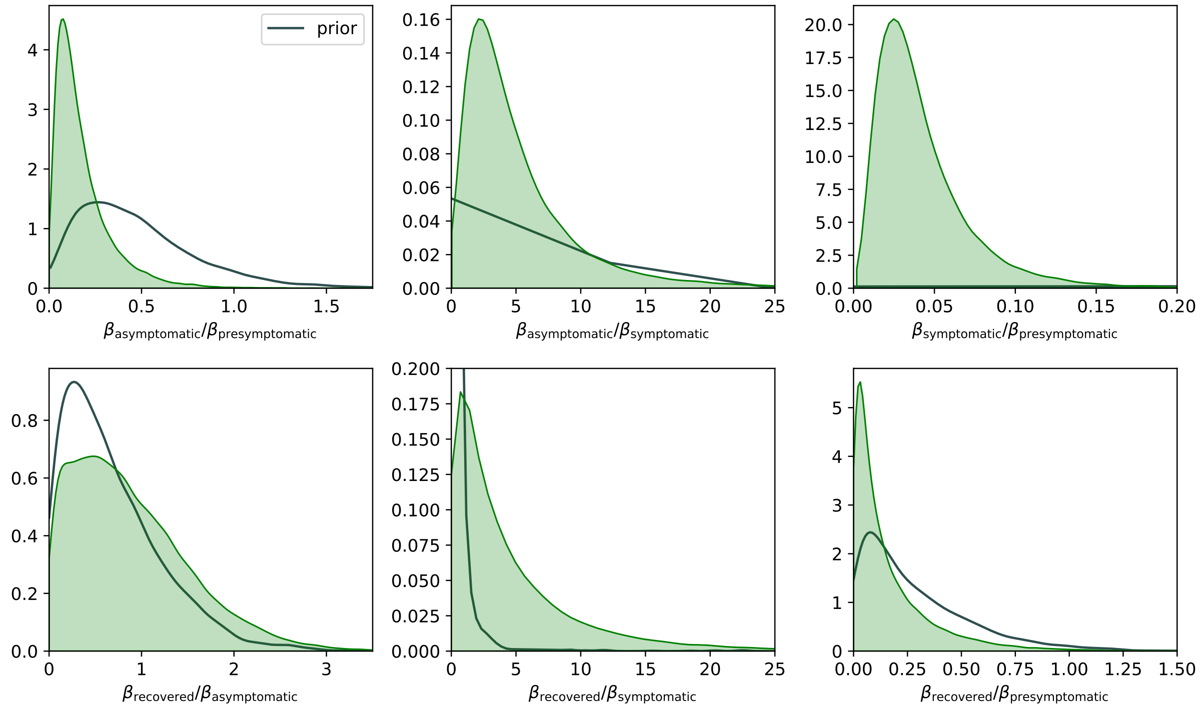


**Supplementary Figure 5: Posterior distributions for the average number $\beta$ of secondary infections per day.** Only the prior and the results obtained using seroprevalence data have been plotted.

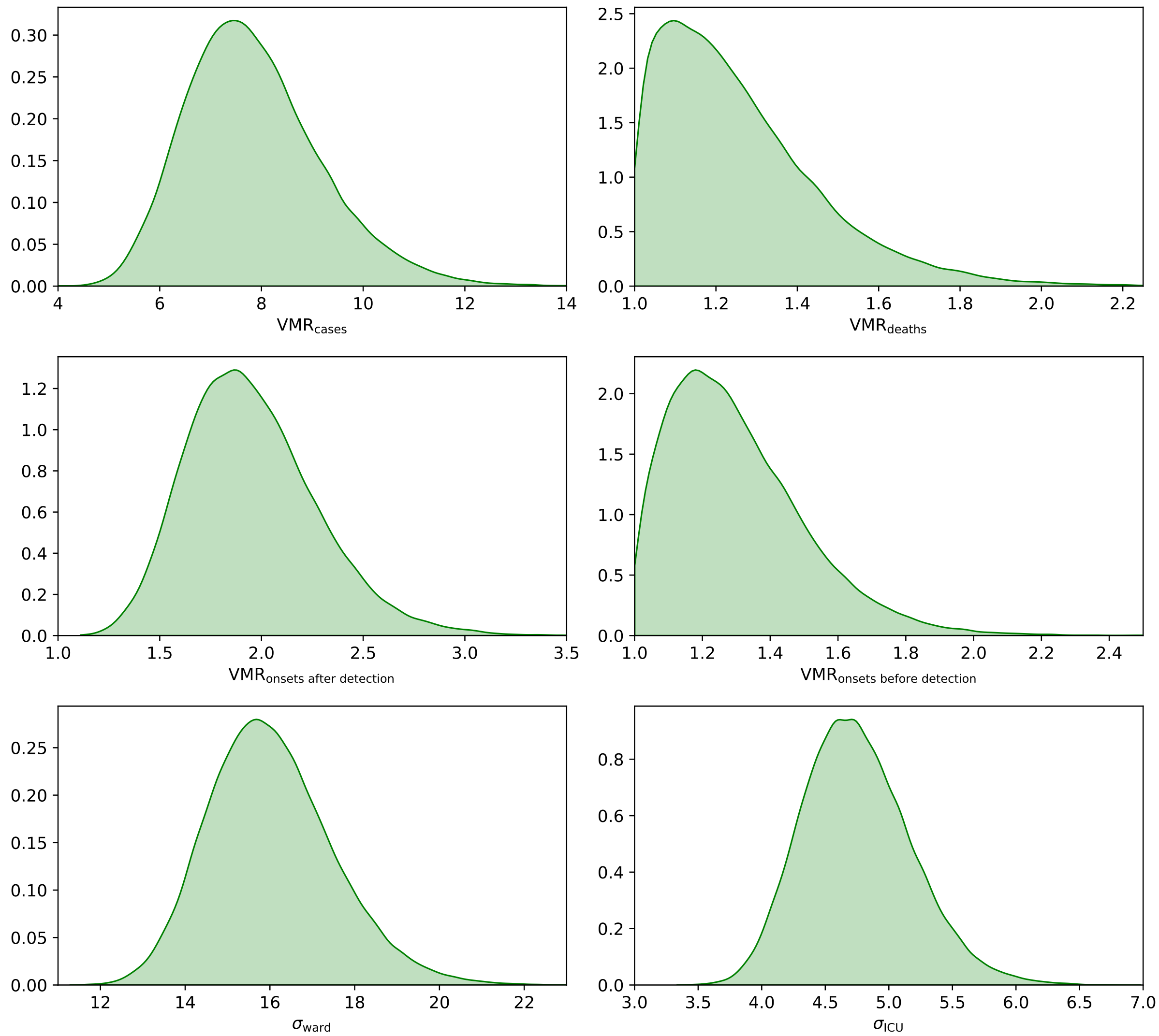


**Supplementary Figure 6: Posterior distributions for the noise levels of the observables.** For negative binomial distributions the variance-to-mean ratio (VMR) is used, while standard deviations $\sigma$ are given for normal distributions. Only the results obtained using seroprevalence data have been plotted.

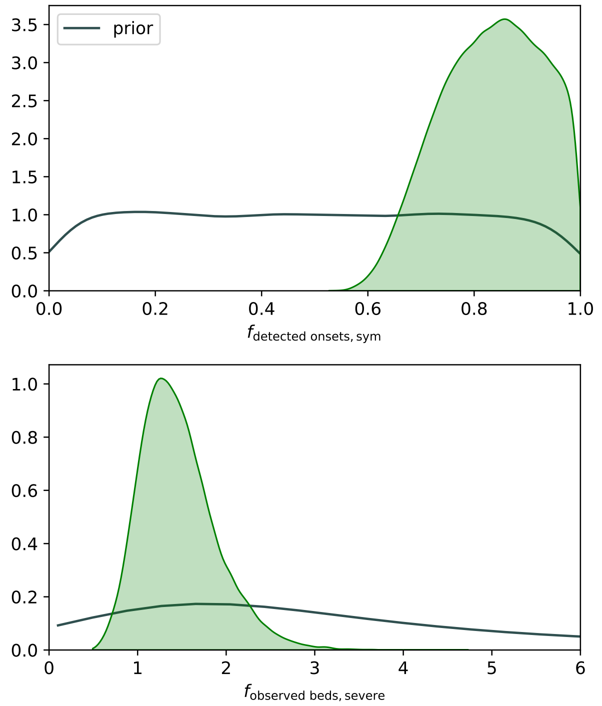


**Supplementary Figure 7: Posterior distributions for the scaling factors of the observables.** The first row shows the scaling factors controlling the probability of a symptom onset date being reported, while the second row shows the scaling factors estimating the under-/over- representation of the observed hospital bed counts due to either data incompleteness or influx of patients from surrounding areas. Only the prior and the results obtained using seroprevalence data have been plotted.

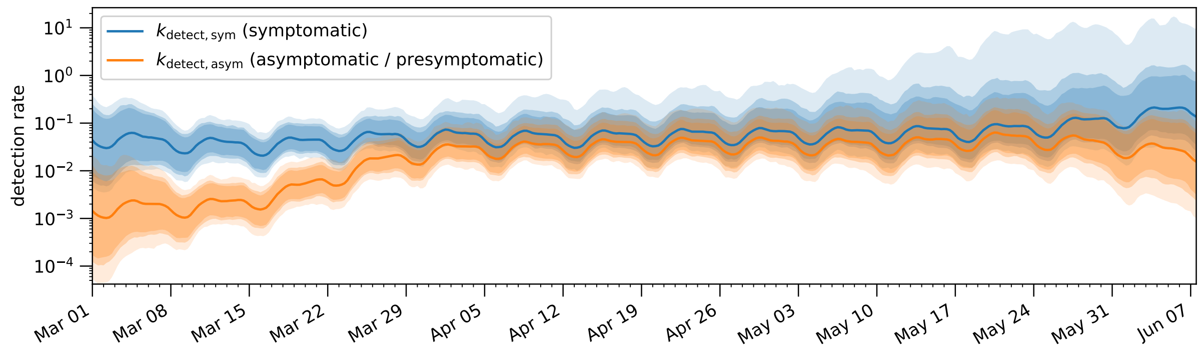


**Supplementary Figure 8: Temporal evolution of the detection rates.** The bands correspond to 85%/90%/95% posterior credible intervals, while the solid line denotes the median value.

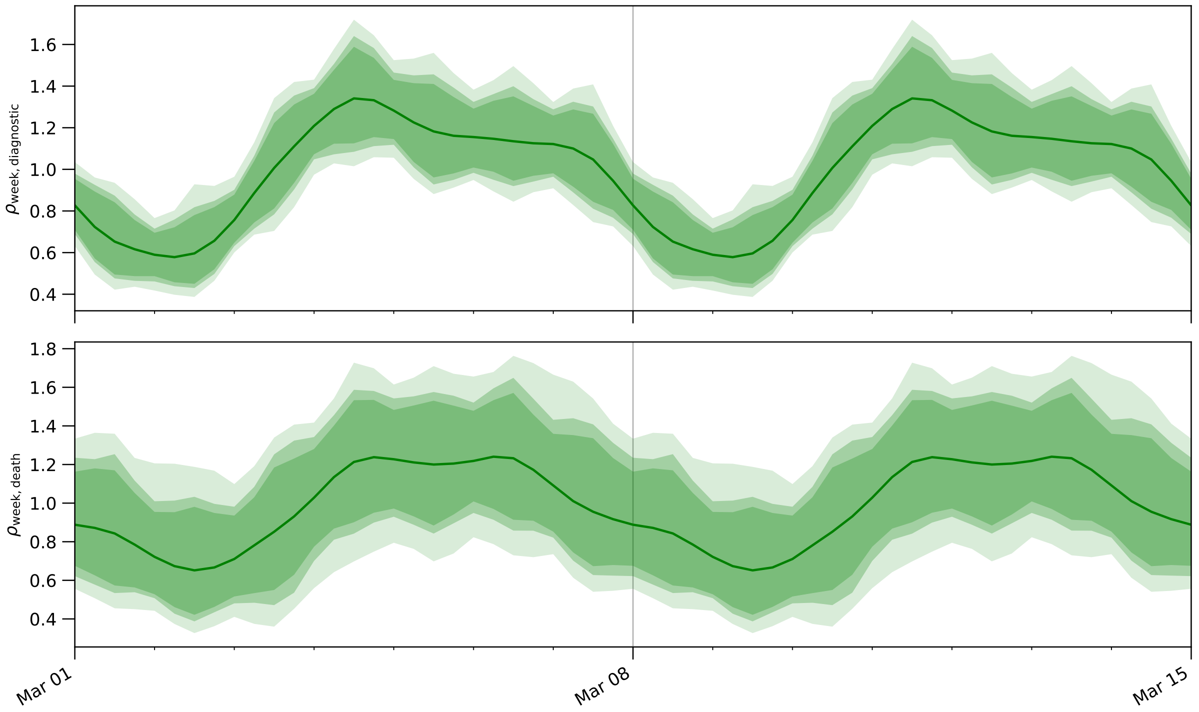


**Supplementary Figure 9: Posterior distributions of the weekly variability in the detection rate and in the death reporting delay.** The bands correspond to 85%/90%/95% posterior credible intervals, while the solid line denotes the median value.

| Transition time (days) | Empirical distribution | Notes | Source | Prior on mean value |
|---|---|---|---|---|
| Incubation | Log$\mathcal{N}(\mu = 1.63, \sigma = 0.5)$<br>mean 5.6, 95% CI 5.0–6.3<br>Erlang$(k = 6, \lambda = 0.88)$ | meta-analysis | McAloon et al. (2020)<br>Linton et al. (2020)<br>Lauer et al. (2020) | Log$\mathcal{N}(\mu = 1.9, \sigma = 0.25)$ |
| Presymptomatic | mean 2.3, 95% CI 0.8–3.0<br>median 2.5, IQR 2–3 | | He et al. (2020)<br>Wei et al. (2020) | $\mathcal{N}(\mu = 2.5, \sigma = 0.85)$ |
| Post-symptomatic infectiousness | mean 13.4 | meta-analysis; hospitalized patients only;<br>detectable virus does not imply infectious | Byrne et al. (2020) | Log$\mathcal{N}(\mu = 2.56, \sigma = 0.35)$ |
| Asymptomatic infectiousness | Gamma(3) with mean 6<br>median 9, IQR 6–11, range 3–21 | | Byrne et al. (2020)<br>Sakurai et al. (2020) | Log$\mathcal{N}(\mu = 2.2, \sigma = 0.35)$ |
| Symptoms | median 7, IQR 3–10 | German study; hospitalized individuals only | Dreher et al. (2020) | $\mathcal{N}(\mu = 7, \sigma = 2.8)$ |
| Symptom onset to hospitalization | mean 7.8, SD 6.2, median 4, IQR 7<br>median 4, IQR 1–8 | international study<br>German study | ISARIC (2020)<br>Dreher et al. (2020) | $\mathcal{N}(\mu = 7.5, \sigma = 2.1)$ |
| Symptom onset to hospitalization (no ICU needed) | median 3, IQR 1–7 | German study;<br>non-ARDS patients (less likely to need ICU) | Dreher et al. (2020) | Log$\mathcal{N}(\mu = 1.57, \sigma = 0.425)$ |
| Symptom onset to hospitalization (ICU needed) | median 7, IQR 2–10 | German study;<br>ARDS patients (more likely to need ICU) | Dreher et al. (2020) | $\mathcal{N}(\mu = 7.5, \sigma = 2.0)$ |
| Symptom onset to ICU | median 9, IQR 4–12 | German study | Dreher et al. (2020) | Log$\mathcal{N}(\mu = 2.14, \sigma = 0.35)$ |
| Hospitalization to ICU | mean 2.9, SD 6.8, median 1, IQR 3 | international study | ISARIC (2020) | $\mathcal{N}(\mu = 2, \sigma = 1.25)$ |
| Hospitalization | mean 12.6, SD 13, median 9, IQR 12<br>median 8, IQR 5–1<br>median 10, IQR 5–19 | international study<br>German study<br>German study;<br>ARDS patients (possible over-estimation) | ISARIC (2020)<br>Dreher et al. (2020)<br>Tolksdorf et al. (2020) | Log$\mathcal{N}(\mu = 2.27, \sigma = 0.265)$ |
| Hospitalization (no ICU needed) | median 7, IQR 5–13 | German study;<br>ARDS patients (possible over-estimation) | Tolksdorf et al. (2020) | $\mathcal{N}(\mu = 7.5, \sigma = 1.575)$ |
| Hospitalization (ICU needed) | median 16, IQR 8–27 | German study;<br>ARDS patients (possible over-estimation) | Tolksdorf et al. (2020) | $\mathcal{N}(\mu = 15.25, \sigma = 4.25)$ |
| Hospitalization (deceased only) | median 10, IQR 4–16 | German study;<br>ARDS patients (more likely to die) | Tolksdorf et al. (2020) | $\mathcal{N}(\mu = 11, \sigma = 4.3)$ |
| ICU stay | mean 13.2, median 8.5, IQR 14.5<br>median 4, IQR 2–10<br>median 5, IQR 2–15 | international study<br>German study<br>German study;<br>ARDS patients (possible over-estimation) | ISARIC (2020)<br>Dreher et al. (2020)<br>Tolksdorf et al. (2020) | Log$\mathcal{N}(\mu = 2.07, \sigma = 0.35)$ |

**Supplementary Table 1:** Prior information on the epidemiological process (transition times).

| Fraction | Empirical estimate | Notes | Source | Prior |
|---|---|---|---|---|
| Mortality (hospitalized individuals) | 22%<br>21% | large-scale German study<br>German study;<br>ARDS patients (more likely to die) | Karagiannidis et al. (2020)<br>Tolksdorf et al. (2020) | $\text{Log}\,\mathcal{N}(\mu = -1.4, \sigma = 0.333)$ |
| Hospitalization | 10%<br>15% | German study | Schilling et al. (2020)<br>Robert Koch Institute (2020) | $\text{Log}\,\mathcal{N}(\mu = -1.96, \sigma = 0.35)$ |
| ICU (hospitalized individuals) | 17%<br>8%<br>17.23%<br><br>13.15%<br><br>37% | international study<br>German study<br>large-scale German study;<br>non-invasive ventilation<br>large-scale German study;<br>invasive ventilation<br>German study;<br>ARDS patients (more likely to need ICU care) | ISARIC (2020)<br>Schilling et al. (2020)<br>Karagiannidis et al. (2020)<br><br>Karagiannidis et al. (2020)<br><br>Tolksdorf et al. (2020) | $\text{Log}\,\mathcal{N}(\mu = -2, \sigma = 0.45)$ |

**Supplementary Table 2:** Prior information on the epidemiological process (fractions).

### References


Byrne, Andrew William et al. (2020). "Inferred duration of infectious period of SARS-CoV-2: rapid scoping review and analysis of available evidence for asymptomatic and symptomatic COVID-19 cases". In: *BMJ Open* 10.8. DOI: 10.1136/bmjopen-2020-039856.

Dreher, Michael et al. (2020). "The Characteristics of 50 Hospitalized COVID-19 Patients With and Without ARDS". In: *Deutsches Ärzteblatt International* 117.16, pp. 271–278. DOI: 10.3238/arztebl.2020.0271.

He, Xi et al. (2020). "Temporal dynamics in viral shedding and transmissibility of COVID-19". In: *Nature Medicine* 26, pp. 672–675. DOI: 10.1038/s41591-020-0869-5.

ISARIC (2020). *COVID-19 Report: 03 September 2020*. URL: https://isaric.org/research/covid-19-clinical-research-resources/evidence-reports/.

Karagiannidis, Christian et al. (2020). "Case characteristics, resource use, and outcomes of 10021 patients with COVID-19 admitted to 920 German hospitals: an observational study". In: 8.9, pp. 853–862. DOI: 10.1016/S2213-2600(20)30316-7.

Lauer, Stephen A. et al. (May 5, 2020). "The Incubation Period of Coronavirus Disease 2019 (COVID-19) From Publicly Reported Confirmed Cases: Estimation and Application". In: *Annals of Internal Medicine* 172.9, pp. 577–582. DOI: 10.7326/M20-0504.

Linton, Natalie M. et al. (2020). "Incubation Period and Other Epidemiological Characteristics of 2019 Novel Coronavirus Infections with Right Truncation: A Statistical Analysis of Publicly Available Case Data". In: *Journal of Clinical Medicine* 9.2. DOI: 10.3390/jcm9020538.

McAloon, Conor et al. (Aug. 2020). "Incubation period of COVID-19: A rapid systematic review and meta-analysis of observational research". In: *BMJ Open* 10 (8), e039652. DOI: 10.1136/BMJOPEN-2020-039652.

Robert Koch Institute (2020). *Epidemiologischer Steckbrief zu SARS-CoV-2 und COVID-19*. URL: https://www.rki.de/DE/Content/InfAZ/N/Neuartiges_Coronavirus/Steckbrief.html.

Sakurai, Aki et al. (2020). "Natural History of Asymptomatic SARS-CoV-2 Infection". In: *New England Journal of Medicine* 383.9, pp. 885–886. DOI: 10.1056/NEJMc2013020.

Schilling, Julia et al. (2020). "Vorläufige Bewertung der Krankheitsschwere von COVID-19 in Deutschland basierend auf übermittelten Fällen gemäß Infektionsschutzgesetz". In: *Epidemiologisches Bulletin* 2020.17, pp. 3–9. DOI: 10.25646/6670.2.

Tolksdorf, Kristin et al. (2020). "Eine höhere Letalität und lange Beatmungsdauer unterscheiden COVID-19 von schwer verlaufenden Atemwegsinfektionen in Grippewellen". In: *Epidemiologisches Bulletin* 41, pp. 3–10. DOI: 10.25646/7111.

Wei, Wycliffe E. et al. (2020). "Presymptomatic Transmission of SARS-CoV-2 – Singapore, January 23-March 16, 2020". In: *Morbidity and Mortality Weekly Report* 69.14, pp. 411–415. DOI: 10.15585/mmwr.mm6914e1.
