## Supplementary material for "Integrative modelling of reported case numbers and seroprevalence reveals time-dependent test efficiency and infectious contacts": Model description and equations

| Symbol | Compartment | Stages for Gamma Chain Trick | Mean transition time |
|---|---|---|---|
| Susc | susceptible individuals | - | (nonlinear) |
| ExpAsym | exposed individuals who will not develop symptoms | 2 | $\tau_{\text{exp}\to\text{asym}}$ |
| ExpSym | exposed individuals who will develop symptoms | 2 | $\tau_{\text{exp}\to\text{sym}}$ |
| Asym | asymptomatic infectious individuals | 3 | $\tau_{\text{asym}}$ |
| ClearAsym | asymptomatic individuals who are no longer infectious | - | (sink state) |
| Pre | pre-symptomatic infectious individuals | 4 | $\tau_{\text{pre}}$ |
| SymMild | symptomatic individuals who will not require hospitalization | 2 | $\tau_{\text{recovery,mild}}$ |
| SymSevere | symptomatic individuals who will require hospitalization but no intensive care | 2 | $\tau_{\text{onset}\to\text{hosp,severe}}$ |
| SymCritical | symptomatic individuals who will require intensive care | 2 | $\tau_{\text{onset}\to\text{hosp,critical}}$ |
| WardSevere | hospitalized individuals who will not require intensive care | 2 | $\tau_{\text{recovery,severe}}$ |
| WardCritical | hospitalized individuals who will require intensive care | 1 | $\tau_{\text{hosp}\to\text{ICU}}$ |
| ICUFatal | individuals in intensive care who will not survive | 1 | $\tau_{\text{ICU}\to\text{death}}$ |
| ICURecoverable | individuals in intensive care who will survive | 1 | $\tau_{\text{recovery,critical}}$ |
| RecMild | individuals from SymMild who are no longer symptomatic but still infectious | 2 | $\tau_{\text{clearance,mild}}$ |
| RecSevere | individuals from SymSevere who are no longer symptomatic but still infectious | 2 | $\tau_{\text{clearance,severe}}$ |
| RecWard | hospitalized individuals who have recovered from intensive care | 1 | $\tau_{\text{discharge}}$ |
| ClearSym | symptomatic individuals who are no longer infectious | - | (sink state) |
| DeadUnreported | deceased individuals who have not yet been reported | 1 | $1/k_{\text{detect,death}}(t)$ |
| DeadReported | deceased individuals who have been reported | - | (sink state) |

**Table 1:** List of model compartments. To each compartment symbol Comp in the table above, there are actually two associated compartments: Comp, containing only the cases not yet reported to the healthcare authorities, and Comp$^*$, containing only cases that have already been reported. Additionally, a numeric superscript denotes the sub-state generated by the Gamma Chain Trick, such as Comp$^{(k)}$ with $k \in \{1, \ldots, \text{number of sub-states}\}$.

| Symbol | Sub-compartments | Description |
|---|---|---|
| Exp | ExpAsym ∪ ExpSym | exposed individuals |
| Clear | ClearSym ∪ ClearAsym | no longer infectious individuals |
| Sym | SymMild ∪ SymSevere ∪ SymCritical | symptomatic (but not hospitalized) individuals |
| Ward | WardSevere ∪ WardCritical ∪ RecWard | hospitalized individuals not in the ICU |
| ICU | ICUFatal ∪ ICURecoverable | individuals in the ICU |
| Hospitalized | Ward ∪ ICU | hospitalized individuals |
| Rec | RecMild ∪ RecSevere | non-hospitalized individuals who are no longer symptomatic |
| Dead | DeadUnreported ∪ DeadReported | deceased individuals |

**Table 2:** List of short-hands for unions of two or more compartments.

| Detection rate | Compartments |
|---|---|
| $k_{\text{detect,asym}}(t)$ | Asym, Pre |
| $k_{\text{detect,sym}}(t)$ | Sym |
| $\infty$ | Hospitalized, Dead |
| 0 | Exp, Rec, Clear |

**Table 3:** Rates at which cases are reported to the healthcare authorities ("detected"). A detection rate equal to $\infty$ means that cases are automatically detected on entry to that compartment; as a consequence only the $^*$ state associated to that compartment exists. A detection rate of 0 does not preclude the existence of an associated $^*$ state, since individuals may have been detected at previous stages.

| $\tilde{\beta}$ | Compartments |
|---|---|
| $\tilde{\beta}_{\text{quarantine}}$ | all $^*$ compartments (detected individuals) |
| $\tilde{\beta}_{\text{sick}}$ | Sym |
| $\tilde{\beta}_{\text{NPI}}(t) \coloneqq \rho_{\text{NPI}}(t)\,\tilde{\beta}_0 + (1-\rho_{\text{NPI}}(t))\,\tilde{\beta}_{\text{sick}}$ | Asym, Pre |

**Table 4:** Average number of potentially infectious contacts $\tilde{\beta}$ that individuals from the various compartments have during one day. The reasonable constraint $\tilde{\beta}_{\text{quarantine}} \leq \tilde{\beta}_{\text{sick}}$ is enforced, while the equally reasonable $\tilde{\beta}_{\text{sick}} \leq \tilde{\beta}_{\text{NPI}}$ holds by construction since $\rho_{\text{NPI}}$ is always positive.

| $\gamma$ | Compartments |
|---|---|
| $\gamma_{\text{asymptomatic}}$ | Asym |
| $\gamma_{\text{presymptomatic}}$ | Pre |
| $\gamma_{\text{symptomatic}}$ | Sym, WardSevere, WardCritical |
| $\gamma_{\text{recovered}}$ | Rec |
| 0 | Exp, ICU |

**Table 5:** Infectiousness level $\gamma$ for each model compartment, i.e., the fraction of potentially infectious contacts $\tilde{\beta}$ which actually result in an infection.

| Parameter | Description |
|---|---|
| $r$ | infection rate |
| $N_0 = 1,561,720$ | initial population |
| $\tilde{\beta}_0 = 7.769$ | average number of potentially infectious contacts an individual has in one day in a pre-pandemic setting |
| $\rho_{\text{NPI}}(t)$ | effectiveness of NPIs at a given timepoint (always positive, 1 corresponds to a pre-pandemic situation and 0 to the most effective NPIs) |
| $f_{\text{asym}}$ | fraction of exposed individuals who do not develop symptoms |
| $f_{\text{hosp}}$ | fraction of symptomatic individuals who are hospitalized |
| $f_{\text{ICU}}$ | fraction of hospitalized individuals who require intensive care |
| $m_{\text{ICU}}$ | fraction of individuals in ICU who do not survive |
| $f_{\text{infected},0}$ | fraction of the total population who is infected with the virus at the simulation start |
| $f_{\text{infectious},0}$ | fraction of infected individuals who are infectious at the simulation start |
| $f_{\text{symptomatic},0}$ | fraction of infectious individuals on the symptomatic model branch who have already had symptoms the simulation start |

**Table 6:** Other parameters.

### ODEs

$$\frac{\mathrm{d}}{\mathrm{d}t}\text{Susc} = -r\,\text{Susc}$$

$$\frac{\mathrm{d}}{\mathrm{d}t}\text{ExpAsym}^{(1)} = f_{\text{asym}} r\,\text{Susc} - \frac{\text{ExpAsym}^{(1)}}{\tau_{\text{exp}\to\text{asym}}/2}$$

$$\frac{\mathrm{d}}{\mathrm{d}t}\text{ExpAsym}^{(2)} = \frac{\text{ExpAsym}^{(1)} - \text{ExpAsym}^{(2)}}{\tau_{\text{exp}\to\text{asym}}/2}$$

$$\frac{\mathrm{d}}{\mathrm{d}t}\text{ExpSym}^{(1)} = (1 - f_{\text{asym}})\,r\,\text{Susc} - \frac{\text{ExpSym}^{(1)}}{\tau_{\text{exp}\to\text{sym}}/2}$$

$$\frac{\mathrm{d}}{\mathrm{d}t}\text{ExpSym}^{(2)} = \frac{\text{ExpSym}^{(1)} - \text{ExpSym}^{(2)}}{\tau_{\text{exp}\to\text{sym}}/2}$$

$$\frac{\mathrm{d}}{\mathrm{d}t}\text{Asym}^{(1)} = \frac{\text{ExpAsym}^{(2)}}{\tau_{\text{exp}\to\text{asym}}/2} - \frac{\text{Asym}^{(1)}}{\tau_{\text{asym}}/3} - k_{\text{detect,asym}}\text{Asym}^{(1)}$$

$$\frac{\mathrm{d}}{\mathrm{d}t}\text{Asym}^{*(1)} = -\frac{\text{Asym}^{*(1)}}{\tau_{\text{asym}}/3} + k_{\text{detect,asym}}\text{Asym}^{(1)}$$

$$\frac{\mathrm{d}}{\mathrm{d}t}\text{Asym}^{(2)} = \frac{\text{Asym}^{(1)} - \text{Asym}^{(2)}}{\tau_{\text{asym}}/3} - k_{\text{detect,asym}}\text{Asym}^{(2)}$$

$$\frac{\mathrm{d}}{\mathrm{d}t}\text{Asym}^{*(2)} = \frac{\text{Asym}^{*(1)} - \text{Asym}^{*(2)}}{\tau_{\text{asym}}/3} + k_{\text{detect,asym}}\text{Asym}^{(2)}$$

$$\frac{\mathrm{d}}{\mathrm{d}t}\text{Asym}^{(3)} = \frac{\text{Asym}^{(2)} - \text{Asym}^{(3)}}{\tau_{\text{asym}}/3} - k_{\text{detect,asym}}\text{Asym}^{(3)}$$

$$\frac{\mathrm{d}}{\mathrm{d}t}\text{Asym}^{*(3)} = \frac{\text{Asym}^{*(2)} - \text{Asym}^{*(3)}}{\tau_{\text{asym}}/3} + k_{\text{detect,asym}}\text{Asym}^{(3)}$$

$$\frac{\mathrm{d}}{\mathrm{d}t}\text{ClearAsym} = \frac{\text{Asym}^{(3)}}{\tau_{\text{asym}}/3}$$

$$\frac{\mathrm{d}}{\mathrm{d}t}\text{ClearAsym}^{*} = \frac{\text{Asym}^{*(3)}}{\tau_{\text{asym}}/3}$$

$$\frac{\mathrm{d}}{\mathrm{d}t}\text{Pre}^{(1)} = \frac{\text{ExpSym}^{(2)}}{\tau_{\text{exp}\to\text{sym}}/2} - \frac{\text{Pre}^{(1)}}{\tau_{\text{pre}}/4} - k_{\text{detect,asym}}\text{Pre}^{(1)}$$

$$\frac{\mathrm{d}}{\mathrm{d}t}\text{Pre}^{*(1)} = -\frac{\text{Pre}^{*(1)}}{\tau_{\text{pre}}/4} + k_{\text{detect,asym}}\text{Pre}^{(1)}$$

$$
\begin{aligned}
\frac{\mathrm{d}}{\mathrm{d}t}\mathrm{Pre}^{(2)} &= \frac{\mathrm{Pre}^{(1)} - \mathrm{Pre}^{(2)}}{\tau_{\mathrm{pre}}/4} - k_{\mathrm{detect,asym}}\mathrm{Pre}^{(2)} \\
\frac{\mathrm{d}}{\mathrm{d}t}\mathrm{Pre}^{*(2)} &= \frac{\mathrm{Pre}^{*(1)} - \mathrm{Pre}^{*(2)}}{\tau_{\mathrm{pre}}/4} + k_{\mathrm{detect,asym}}\mathrm{Pre}^{(2)} \\
\frac{\mathrm{d}}{\mathrm{d}t}\mathrm{Pre}^{(3)} &= \frac{\mathrm{Pre}^{(2)} - \mathrm{Pre}^{(3)}}{\tau_{\mathrm{pre}}/4} - k_{\mathrm{detect,asym}}\mathrm{Pre}^{(3)} \\
\frac{\mathrm{d}}{\mathrm{d}t}\mathrm{Pre}^{*(3)} &= \frac{\mathrm{Pre}^{*(2)} - \mathrm{Pre}^{*(3)}}{\tau_{\mathrm{pre}}/4} + k_{\mathrm{detect,asym}}\mathrm{Pre}^{(3)} \\
\frac{\mathrm{d}}{\mathrm{d}t}\mathrm{Pre}^{(4)} &= \frac{\mathrm{Pre}^{(3)} - \mathrm{Pre}^{(4)}}{\tau_{\mathrm{pre}}/4} - k_{\mathrm{detect,asym}}\mathrm{Pre}^{(4)} \\
\frac{\mathrm{d}}{\mathrm{d}t}\mathrm{Pre}^{*(4)} &= \frac{\mathrm{Pre}^{*(3)} - \mathrm{Pre}^{*(4)}}{\tau_{\mathrm{pre}}/4} + k_{\mathrm{detect,asym}}\mathrm{Pre}^{(4)} \\
\frac{\mathrm{d}}{\mathrm{d}t}\mathrm{SymMild}^{(1)} &= (1 - f_{\mathrm{hosp}})\frac{\mathrm{Pre}^{(4)}}{\tau_{\mathrm{pre}}/4} - \frac{\mathrm{SymMild}^{(1)}}{\tau_{\mathrm{recovery,mild}}/2} - k_{\mathrm{detect,sym}}\mathrm{SymMild}^{(1)} \\
\frac{\mathrm{d}}{\mathrm{d}t}\mathrm{SymMild}^{*(1)} &= (1 - f_{\mathrm{hosp}})\frac{\mathrm{Pre}^{*(4)}}{\tau_{\mathrm{pre}}/4} - \frac{\mathrm{SymMild}^{*(1)}}{\tau_{\mathrm{recovery,mild}}/2} + k_{\mathrm{detect,sym}}\mathrm{SymMild}^{(1)} \\
\frac{\mathrm{d}}{\mathrm{d}t}\mathrm{SymMild}^{(2)} &= \frac{\mathrm{SymMild}^{(1)} - \mathrm{SymMild}^{(2)}}{\tau_{\mathrm{recovery,mild}}/2} - k_{\mathrm{detect,sym}}\mathrm{SymMild}^{(2)} \\
\frac{\mathrm{d}}{\mathrm{d}t}\mathrm{SymMild}^{*(2)} &= \frac{\mathrm{SymMild}^{*(1)} - \mathrm{SymMild}^{*(2)}}{\tau_{\mathrm{recovery,mild}}/2} + k_{\mathrm{detect,sym}}\mathrm{SymMild}^{(2)} \\
\frac{\mathrm{d}}{\mathrm{d}t}\mathrm{SymSevere} &= (1 - f_{\mathrm{ICU}})\, f_{\mathrm{hosp}}\frac{\mathrm{Pre}^{(4)}}{\tau_{\mathrm{pre}}/4} - \frac{\mathrm{SymSevere}}{\tau_{\mathrm{onset}\to\mathrm{hosp,severe}}} - k_{\mathrm{detect,sym}}\mathrm{SymSevere} \\
\frac{\mathrm{d}}{\mathrm{d}t}\mathrm{SymSevere}^{*} &= (1 - f_{\mathrm{ICU}})\, f_{\mathrm{hosp}}\frac{\mathrm{Pre}^{*(4)}}{\tau_{\mathrm{pre}}/4} - \frac{\mathrm{SymSevere}^{*}}{\tau_{\mathrm{onset}\to\mathrm{hosp,severe}}} + k_{\mathrm{detect,sym}}\mathrm{SymSevere} \\
\frac{\mathrm{d}}{\mathrm{d}t}\mathrm{SymCritical}^{(1)} &= f_{\mathrm{ICU}} f_{\mathrm{hosp}}\frac{\mathrm{Pre}^{(4)}}{\tau_{\mathrm{pre}}/4} - \frac{\mathrm{SymCritical}^{(1)}}{\tau_{\mathrm{onset}\to\mathrm{hosp,critical}}/2} - k_{\mathrm{detect,sym}}\mathrm{SymCritical}^{(1)} \\
\frac{\mathrm{d}}{\mathrm{d}t}\mathrm{SymCritical}^{*(1)} &= f_{\mathrm{ICU}} f_{\mathrm{hosp}}\frac{\mathrm{Pre}^{*(4)}}{\tau_{\mathrm{pre}}/4} - \frac{\mathrm{SymCritical}^{*(1)}}{\tau_{\mathrm{onset}\to\mathrm{hosp,critical}}/2} + k_{\mathrm{detect,sym}}\mathrm{SymCritical}^{(1)} \\
\frac{\mathrm{d}}{\mathrm{d}t}\mathrm{SymCritical}^{(2)} &= \frac{\mathrm{SymCritical}^{(1)} - \mathrm{SymCritical}^{(2)}}{\tau_{\mathrm{onset}\to\mathrm{hosp,critical}}/2} - k_{\mathrm{detect,sym}}\mathrm{SymCritical}^{(2)} \\
\frac{\mathrm{d}}{\mathrm{d}t}\mathrm{SymCritical}^{*(2)} &= \frac{\mathrm{SymCritical}^{*(1)} - \mathrm{SymCritical}^{*(2)}}{\tau_{\mathrm{onset}\to\mathrm{hosp,critical}}/2} + k_{\mathrm{detect,sym}}\mathrm{SymCritical}^{(2)} \\
\frac{\mathrm{d}}{\mathrm{d}t}\mathrm{WardSevere}^{*(1)} &= \frac{\mathrm{SymSevere} + \mathrm{SymSevere}^{*}}{\tau_{\mathrm{onset}\to\mathrm{hosp,severe}}} - \frac{\mathrm{WardSevere}^{*(1)}}{\tau_{\mathrm{ward}\to\mathrm{recovered}}/2} \\
\frac{\mathrm{d}}{\mathrm{d}t}\mathrm{WardSevere}^{*(2)} &= \frac{\mathrm{WardSevere}^{*(1)} - \mathrm{WardSevere}^{*(2)}}{\tau_{\mathrm{ward}\to\mathrm{recovered}}/2} \\
\frac{\mathrm{d}}{\mathrm{d}t}\mathrm{WardCritical}^{*} &= \frac{\mathrm{SymCritical}^{(2)} + \mathrm{SymCritical}^{*(2)}}{\tau_{\mathrm{onset}\to\mathrm{hosp,critical}}/2} - \frac{\mathrm{WardCritical}^{*}}{\tau_{\mathrm{hosp}\to\mathrm{ICU}}} \\
\frac{\mathrm{d}}{\mathrm{d}t}\mathrm{ICUFatal}^{*} &= m_{\mathrm{ICU}}\frac{\mathrm{WardCritical}^{*}}{\tau_{\mathrm{hosp}\to\mathrm{ICU}}} - \frac{\mathrm{ICUFatal}^{*}}{\tau_{\mathrm{ICU}\to\mathrm{death}}} \\
\frac{\mathrm{d}}{\mathrm{d}t}\mathrm{ICURecoverable}^{*} &= (1 - m_{\mathrm{ICU}})\frac{\mathrm{WardCritical}^{*}}{\tau_{\mathrm{hosp}\to\mathrm{ICU}}} - \frac{\mathrm{ICURecoverable}^{*}}{\tau_{\mathrm{recovery,critical}}} \\
\frac{\mathrm{d}}{\mathrm{d}t}\mathrm{RecMild}^{(1)} &= \frac{\mathrm{SymMild}^{(2)}}{\tau_{\mathrm{recovery,mild}}/2} - \frac{\mathrm{RecMild}^{(1)}}{\tau_{\mathrm{clearance,mild}}/2} \\
\frac{\mathrm{d}}{\mathrm{d}t}\mathrm{RecMild}^{*(1)} &= \frac{\mathrm{SymMild}^{*(2)}}{\tau_{\mathrm{recovery,mild}}/2} - \frac{\mathrm{RecMild}^{*(1)}}{\tau_{\mathrm{clearance,mild}}/2} \\
\frac{\mathrm{d}}{\mathrm{d}t}\mathrm{RecMild}^{(2)} &= \frac{\mathrm{RecMild}^{(1)} - \mathrm{RecMild}^{(2)}}{\tau_{\mathrm{clearance,mild}}/2} \\
\frac{\mathrm{d}}{\mathrm{d}t}\mathrm{RecMild}^{*(2)} &= \frac{\mathrm{RecMild}^{*(1)} - \mathrm{RecMild}^{*(2)}}{\tau_{\mathrm{clearance,mild}}/2}
\end{aligned}
$$

$$
\begin{aligned}
\frac{\mathrm{d}}{\mathrm{d}t}\mathrm{RecSevere}^{*(1)} &= \frac{\mathrm{WardSevere}^{*(2)}}{\tau_{\mathrm{ward}\to\mathrm{recovered}}/2} - \frac{\mathrm{RecSevere}^{*(1)}}{\tau_{\mathrm{clearance,severe}}/2} \\
\frac{\mathrm{d}}{\mathrm{d}t}\mathrm{RecSevere}^{*(2)} &= \frac{\mathrm{RecSevere}^{*(1)} - \mathrm{RecSevere}^{*(2)}}{\tau_{\mathrm{clearance,severe}}/2} \\
\frac{\mathrm{d}}{\mathrm{d}t}\mathrm{RecWard}^{*} &= \frac{\mathrm{ICURecoverable}^{*}}{\tau_{\mathrm{recovery,critical}}} - \frac{\mathrm{RecWard}^{*}}{\tau_{\mathrm{discharge}}} \\
\frac{\mathrm{d}}{\mathrm{d}t}\mathrm{ClearSym} &= \frac{\mathrm{RecMild}^{(2)}}{\tau_{\mathrm{clearance,mild}}/2} \\
\frac{\mathrm{d}}{\mathrm{d}t}\mathrm{ClearSym}^{*} &= \frac{\mathrm{RecMild}^{*(2)}}{\tau_{\mathrm{clearance,mild}}/2} + \frac{\mathrm{RecSevere}^{*(2)}}{\tau_{\mathrm{clearance,severe}}/2} + \frac{\mathrm{RecWard}^{*}}{\tau_{\mathrm{discharge}}} \\
\frac{\mathrm{d}}{\mathrm{d}t}\mathrm{DeadUnreported}^{*} &= \frac{\mathrm{ICUFatal}^{*}}{\tau_{\mathrm{ICU}\to\mathrm{death}}} - k_{\mathrm{detect,death}}\mathrm{DeadUnreported}^{*} \\
\frac{\mathrm{d}}{\mathrm{d}t}\mathrm{DeadReported}^{*} &= k_{\mathrm{detect,death}}\mathrm{DeadUnreported}^{*}
\end{aligned}
$$

### Infection rate

$$
\begin{aligned}
N_0 r = \quad & \tilde{\beta}_{\mathrm{NPI}}\gamma_{\mathrm{asymptomatic}}\mathrm{Asym} \\
& + \tilde{\beta}_{\mathrm{NPI}}\gamma_{\mathrm{presymptomatic}}\mathrm{Pre} \\
& + \tilde{\beta}_{\mathrm{sick}}\gamma_{\mathrm{symptomatic}}\left(\mathrm{SymCritical} + \mathrm{SymMild} + \mathrm{SymSevere}\right) \\
& + \tilde{\beta}_{\mathrm{NPI}}\gamma_{\mathrm{recovered}}\mathrm{RecMild} \\
& + \tilde{\beta}_{\mathrm{quarantine}}\gamma_{\mathrm{asymptomatic}}\mathrm{Asym}^{*} \\
& + \tilde{\beta}_{\mathrm{quarantine}}\gamma_{\mathrm{presymptomatic}}\mathrm{Pre}^{*} \\
& + \tilde{\beta}_{\mathrm{quarantine}}\gamma_{\mathrm{symptomatic}}\left(\mathrm{SymCritical}^{*} + \mathrm{SymMild}^{*} + \mathrm{SymSevere}^{*} + \mathrm{WardCritical}^{*} + \mathrm{WardSevere}^{*}\right) \\
& + \tilde{\beta}_{\mathrm{quarantine}}\gamma_{\mathrm{recovered}}\left(\mathrm{RecMild}^{*} + \mathrm{RecSevere}^{*}\right)
\end{aligned}
$$

### Initial conditions

$$
\begin{aligned}
\mathrm{Susc}(0) &= \left(1 - f_{\mathrm{infected},0}\right) N_0 \\
\mathrm{ExpAsym}^{(i)}(0) &= \frac{1}{2} f_{\mathrm{asym}}\left(1 - f_{\mathrm{infectious},0}\right) f_{\mathrm{infected},0} N_0, \quad i = 1,2 \\
\mathrm{ExpSym}^{(i)}(0) &= \frac{1}{2}\left(1 - f_{\mathrm{asym}}\right)\left(1 - f_{\mathrm{infectious},0}\right) f_{\mathrm{infected},0}\, N_0, \quad i = 1,2 \\
\mathrm{Asym}^{(i)}(0) &= \frac{1}{3} f_{\mathrm{asym}}\, f_{\mathrm{infectious},0}\, f_{\mathrm{infected},0}\, N_0, \quad i = 1,2,3 \\
\mathrm{Pre}^{(i)}(0) &= \frac{1}{4}\left(1 - f_{\mathrm{symptomatic},0}\right)\left(1 - f_{\mathrm{asym}}\right) f_{\mathrm{infectious},0}\, f_{\mathrm{infected},0}\, N_0, \quad i = 1,2,3,4 \\
\mathrm{SymMild}^{(i)}(0) &= \frac{1}{2}\left(1 - f_{\mathrm{hosp}}\right) f_{\mathrm{symptomatic},0}\left(1 - f_{\mathrm{asym}}\right) f_{\mathrm{infectious},0}\, f_{\mathrm{infected},0}\, N_0, \quad i = 1,2 \\
\mathrm{SymSevere}(0) &= \left(1 - f_{\mathrm{ICU}}\right) f_{\mathrm{hosp}}\, f_{\mathrm{symptomatic},0}\left(1 - f_{\mathrm{asym}}\right) f_{\mathrm{infectious},0}\, f_{\mathrm{infected},0}\, N_0 \\
\mathrm{SymCritical}^{(i)}(0) &= \frac{1}{2} f_{\mathrm{ICU}}\, f_{\mathrm{hosp}}\, f_{\mathrm{symptomatic},0}\left(1 - f_{\mathrm{asym}}\right) f_{\mathrm{infectious},0}\, f_{\mathrm{infected},0}\, N_0, \quad i = 1,2
\end{aligned}
$$

$$
\text{all other compartments, in particular all } {}^{*} \text{ compartments} = 0
$$

#### Observables

**Rate at which new cases are reported to the healthcare authorities**

$$\frac{\mathrm{SymSevere}}{\tau_{\mathrm{onset}\to\mathrm{hosp,severe}}} + \frac{\mathrm{SymCritical}^{(2)}}{\tau_{\mathrm{onset}\to\mathrm{hosp,critical}}/2} + k_{\mathrm{detect,asym}}\left(\mathrm{Asym} + \mathrm{Pre}\right) + k_{\mathrm{detect,sym}}\mathrm{Sym}$$

**Rate at which deaths are reported to the healthcare authorities**

$$k_{\mathrm{detect,death}}\mathrm{DeadUnreported}^{*}$$

**Rate at which symptom onsets are reported to the healthcare authorities (by individuals who have been detected after the onset of symptoms)**

$$p_{\mathrm{detected}}\frac{\mathrm{Pre}^{(4)}}{\tau_{\mathrm{pre}}/4}$$

where $p_{\mathrm{detected}}$ is the probability that a person who is showing symptoms for the first time and has not been detected yet will be detected before the virus is cleared from their system, as given by the formula

$$p_{\mathrm{detected}} = f_{\mathrm{detected\ onsets,sym}}\left[1 - \left(1 - f_{\mathrm{hosp}}\right)\left(\frac{\tau_{\mathrm{test,sym,0}}}{\tau_{\mathrm{recovery,mild}}/2 + \tau_{\mathrm{test,sym,0}}}\right)^{2}\right]$$

**Rate at which symptom onsets are reported to the healthcare authorities (by individuals who have been detected before the onset of symptoms, i.e., while presymptomatic)**

$$f_{\mathrm{detected\ onsets,asym}}\frac{\mathrm{Pre}^{*(4)}}{\tau_{\mathrm{pre}}/4}$$

**Number of hospital ward (not ICU) beds occupied by COVID-19 patients**

$$f_{\mathrm{observed\ beds,severe}}\mathrm{WardSevere}^{*} + f_{\mathrm{observed\ beds,critical}}\left(\mathrm{RecWard}^{*} + \mathrm{WardCritical}^{*}\right)$$

**Number of hospital ICU beds occupied by COVID-19 patients**

$$f_{\mathrm{observed\ beds,critical}}\mathrm{ICU}^{*}$$

**Measured prevalence of COVID-19 antibodies in the population at time $t$**

$$(\mathrm{se} + \mathrm{sp} - 1)\frac{\hat{N}(t - 2\,\mathrm{weeks})}{N_0} + (1 - \mathrm{sp})$$

where se = 0.8860104 and sp = 0.9972041 are the sensitivity and specificity of the antibody test, while $\hat{N}(t)$ is the number of individuals who will possess COVID-19 antibodies two weeks after the time point $t$, as given by

$$\begin{aligned}\hat{N} = \quad & \mathrm{Asym} + \mathrm{Asym}^{*} + \mathrm{SymMild} + \mathrm{SymMild}^{*} + \mathrm{SymSevere} + \mathrm{SymSevere}^{*} \\ & + \mathrm{WardSevere}^{*} + \mathrm{ICURecoverable}^{*} + \mathrm{Rec} + \mathrm{Rec}^{*} + \mathrm{RecWard}^{*} + \mathrm{Clear} + \mathrm{Clear}^{*} \\ & + \left(1 - m_{\mathrm{ICU}}\right)\left(\mathrm{SymCritical} + \mathrm{SymCritical}^{*} + \mathrm{WardCritical}^{*}\right) + \left(1 - m_{\mathrm{ICU}}f_{\mathrm{ICU}}f_{\mathrm{hosp}}\right)\left(\mathrm{Pre} + \mathrm{Pre}^{*}\right)\end{aligned}$$
